# Three-dimensional motion-capture of the heart uncovers signatures of human health and disease

**DOI:** 10.1101/2025.05.22.25328000

**Authors:** Pierre-Raphael Schiratti, Soodeh Kalaie, Jin Zheng, Paolo Inglese, Kathryn A McGurk, Sean L Zheng, James S Ware, Wenjia Bai, Declan P O’Regan

## Abstract

Adaptations of the motion dynamics of the heart are key to understanding pathophysiological mechanisms and transitions from health to disease. Conventional volumetric assessments of the heart using imaging represent mainly aggregate global features of function that are poorly discriminating. Here we aimed to optimise understanding of how the heart is affected by cardiovascular risk factors through efficient representations of motion trajectories. We use computer vision to survey three-dimensional cardiac motion traits using densely sampled point clouds of the heart in over 20,000 participants of UK Biobank. We developed a computational framework for dimensionality reduction of spatiotemporal information to derive a human-interpretable signature summarising variation in complex patterns of motion. We found six phenogroups representing a novel classification of heterogenous motion phenotypes with differential enrichment of cardiovascular outcomes and genetic risk. Low dimensional representations of motion are visualised as a simple spatial signature capturing deviation from an average state. Discovering compact cardiac motion signatures of health and disease enables efficient classification of patient risk and predisposing polygenic factors.

## Introduction

The motion dynamics of the beating heart comprise a complex rhythmic pattern of non-linear trajectories regulated by molecular, electrical and biophysical processes that are susceptible to disease.^1^ Conventional approaches for assessing cardiovascular function have relied on simple volumetric measures obtained using imaging however such approaches have significant limitations as they capture only composite global markers of function that are insensitive to complex traits of contractile coordination, fibre geometry and active relaxation.^2^ The measurement and interpretation of conventional parameters also relies on expertise and is constrained by one-dimensional dichotomous thresholds for diagnosis.^3^ Cardiomyopathies for instance are characterised by complex regional patterns of remodelling and early signs of disease may not be apparent using only volumetric parameters.^4^

Computer vision approaches now enable three-dimensional cardiac motion estimation from sparse medical imaging of the heart. These approaches transform planar cross-sectional imaging into a digital representation of whole heart motion that can be aggregated across large and diverse populations.^5,6^ While rich representations of motion contain latent prognostic information,^1^ their value in classifying disease sub-types is not yet known. Unsupervised deep learning approaches can efficiently classify patients from controls using complex traits,^7^ but are limited to static geometries and do not provide interpretable features. Here, we present a novel framework for characterising diversity in cardiac motion using spatio-temporal point clouds derived from cardiac magnetic resonance (CMR) imaging. We developed a convolutional variational autoencoder to embed these complex cardiac models in a low-dimensional latent space. Temporal signatures were clustered on latent features of motion to define six phenogroups with differential enrichment of cardiovascular risk factors, polygenic scores and clinical outcomes. A human interpretable signature allows visualisation of how each three dimensional motion state deviates from a normative reference.

## Results

### Study overview

The UK Biobank study recruited 500,000 participants aged 40 to 69 years between 2006 and 2010.^8^ Genotyping array and exome sequencing were available for over 450,000 participants. A sub-study recalled participants for CMR,^9^ which included retrospectively-gated short axis cines, and here we analysed data from 20,318 participants. Distributions of demographic and image-derived features in the population are reported in Table 1. A flowchart (Figure 1) describes each analysis step. Computer vision techniques transformed the two dimensional cine images of the heart into a time-resolved three-dimensional point cloud of the endo and epicardial surfaces of the heart (Figure 2). Each model of the left ventricle comprised 40,616 points (downsampled to 1406) over 50 time frames throughout the cardiac cycle. Atlas registration ensured that the number and position of points was consistent between participants as well as at each cardiac phase.

**Table 1.** Description of UK Biobank participants. Principal characteristics of UK Biobank participants with motion cardiac imaging data stratified by sex. Data on demographics, clinical measurements and MRI-derived measures were available. Statistics are described in total count and frequency in brackets for discrete features or mean and standard deviation in brackets for continuous features. mmHg, millimetre of mercury; CMR, cardiac magnetic resonance; LV, left ventricle; RV, right ventricle.

| Trait | Overall | Female | Male |
| --- | --- | --- | --- |
| n | 20318 | 10695 | 9623 |
| Age (years) | 63.0 (7.6) | 62.4 (7.5) | 63.7 (7.7) |
| Body surface area (m<sup>2</sup>) | 1.9 (0.2) | 1.7 (0.2) | 2.0 (0.2) |
| Ethnic background |  |  |  |
| British | 18565 (91.4) | 9769 (91.3) | 8796 (91.4) |
| Any other white background | 557 (2.7) | 323 (3.0) | 234 (2.4) |
| Irish | 551 (2.7) | 290 (2.7) | 261 (2.7) |
| Indian | 147 (0.7) | 56 (0.5) | 91 (0.9) |
| Other ethnic group | 90 (0.4) | 47 (0.4) | 43 (0.4) |
| Diastolic blood pressure (mmHg) | 78.7 (10.5) | 77.0 (10.5) | 80.5 (10.2) |
| Systolic blood pressure (mmHg) | 139.4 (19.4) | 136.3 (20.1) | 142.7 (18.0) |
| MRI measures |  |  |  |
| Circumferential strain (%) | -22.3 (3.4) | -23.4 (3.1) | -21.1 (3.2) |
| Radial strain (%) | 45.0 (8.3) | 47.5 (7.9) | 42.2 (7.8) |
| Longitudinal strain (%) | -18.5 (2.8) | -19.1 (2.7) | -17.8 (2.7) |
| LV end-diastolic volume (mL) | 147.9 (33.5) | 129.4 (22.5) | 168.4 (31.6) |
| LV end-systolic volume (mL) | 60.4 (19.0) | 50.5 (12.7) | 71.4 (18.9) |
| LV ejection fraction (%) | 59.6 (6.1) | 61.1 (5.6) | 57.8 (6.1) |
| LV mass (mg) | 86.0 (22.1) | 70.9 (12.3) | 102.7 (18.3) |
| LV maximum wall thickness (mm) | 9.4 (1.8) | 8.6 (1.4) | 10.3 (1.7) |
| RV end-diastolic volume (mL) | 156.5 (37.3) | 134.2 (24.2) | 181.3 (33.4) |
| RV systolic volume (mL) | 67.6 (21.3) | 54.9 (13.3) | 81.6 (19.5) |
| RV ejection fraction (%) | 57.3 (6.2) | 59.2 (5.7) | 55.1 (5.9) |

**Figure 1.**
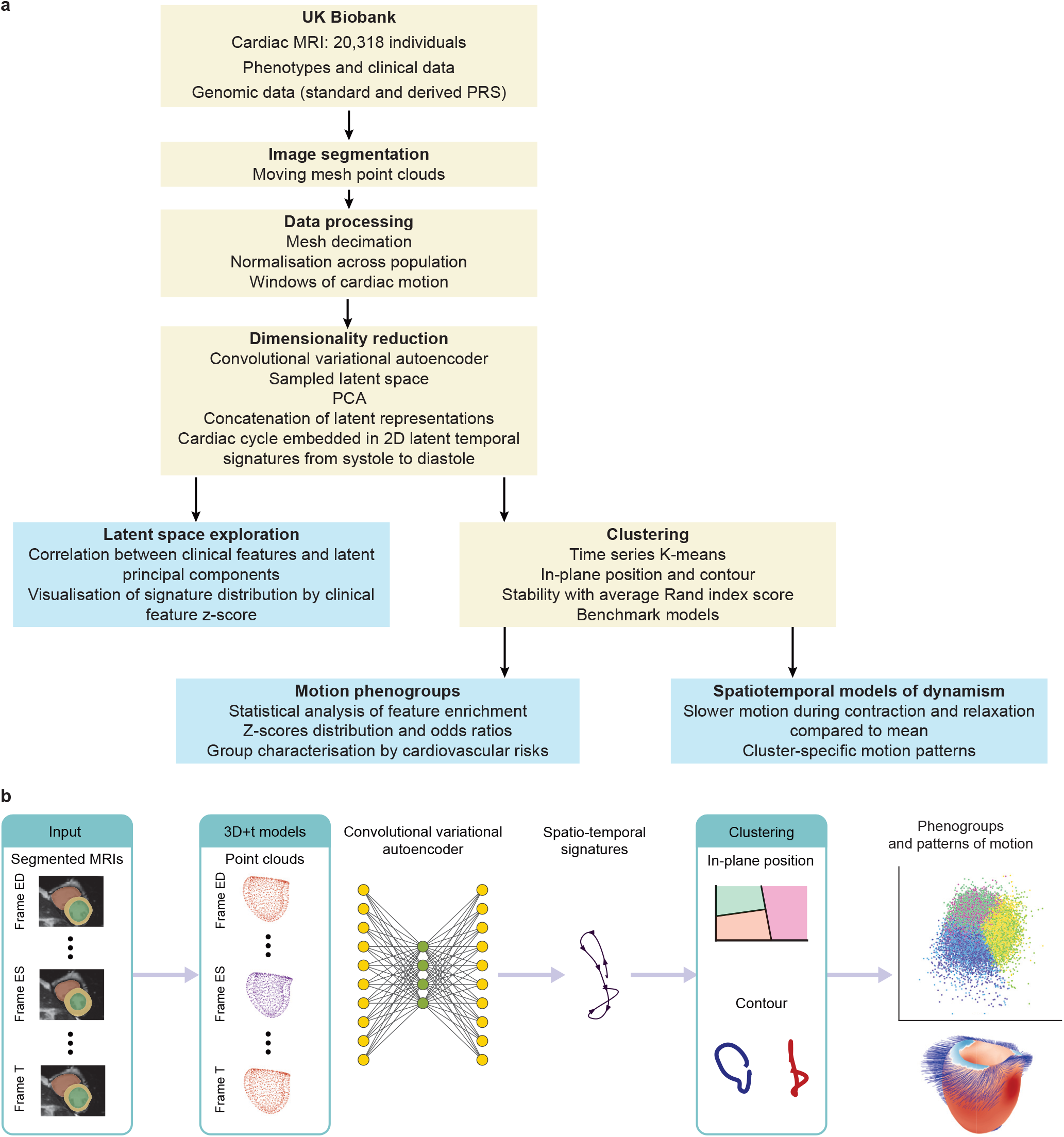
Flowchart of the analysis. **a**. Main steps of the generation of cardiac motion signatures and analysis of clinically relevant novel phenogroups. **b**. Outline of the transformation of cardiac motion data into human-interpretable temporal signatures. Cardiovascular risk groupings and their associated cardiac motion patterns were derived from these signatures. MRI, magnetic resonance imaging; PRS, polygenic risk score; PCA, principal component analysis.

**Figure 2.**
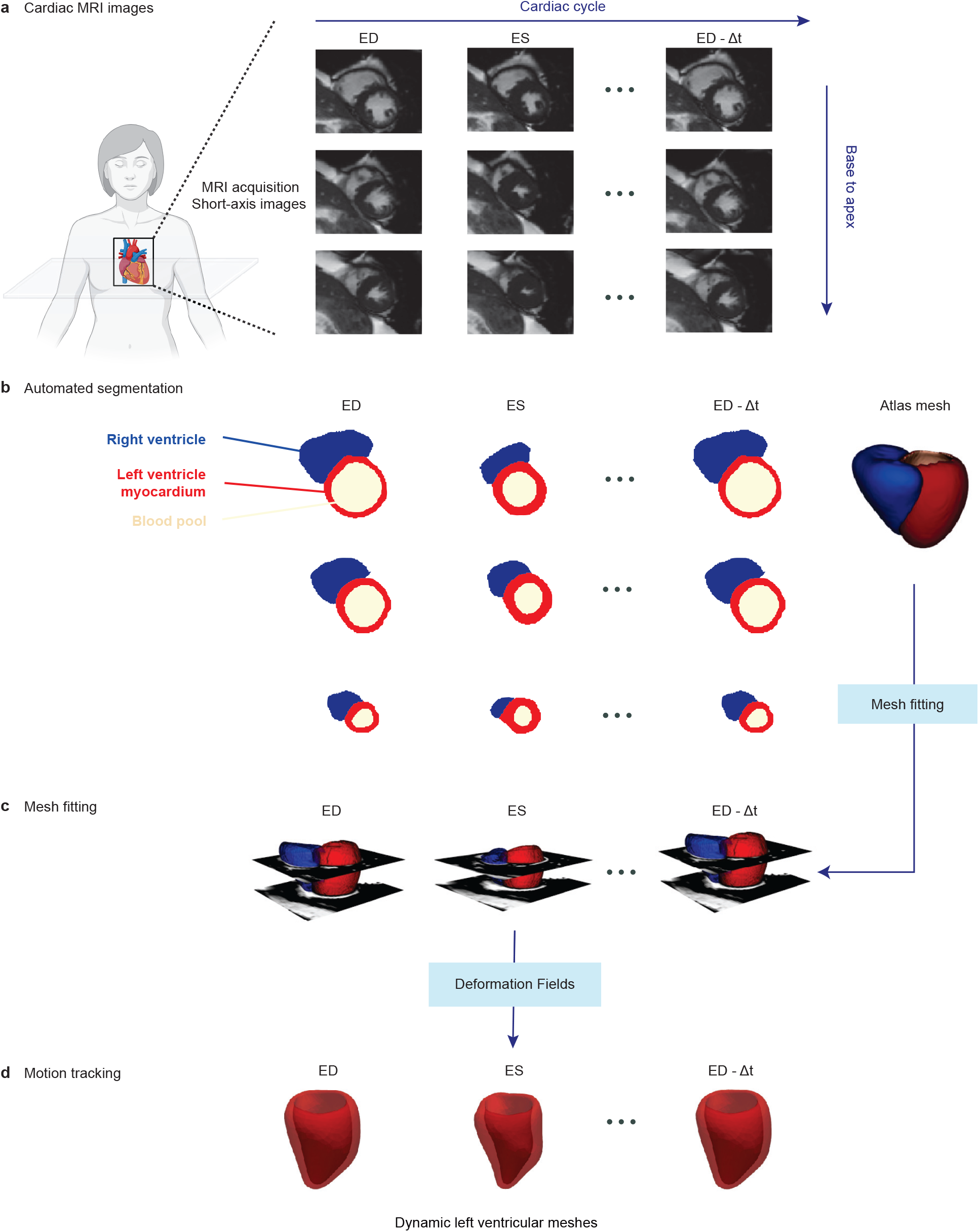
Cardiac imaging segmentation and spatio-temporal models. **a**. Cardiac MRI images were acquired for each participant. They capture the anatomy of the heart from base to apex of ventricles and the temporal data throughout the cardiac cycle from diastole to systole. **b**. These images were then automatically segmented with our algorithm to locate the right ventricle and the left ventricle myocardium and blood pool. **c**. These segmentation masks were then fitted and registered to a common atlas space between participants to develop a consistent three-dimensional model of the heart. **d**. Deformation fields were used to generate the entire cardiac cycle of surface meshes.

To capture spatio-temporal patterns throughout the cardiac cycle, sequences of consecutive point cloud frames were constructed using an overlapping sliding window approach. Dimensionality reduction was performed using a convolutional variational autoencoder followed by a principal component analysis (PCA). Clustering of spatio-temporal patterns was performed with K-means for time series data. Six clusters (phenogroups) of motion traits were summarised by enrichment for prevalent disease, future events and polygenic risk scores. Each phenogroup was compared to the population average to define a visually interpretable signature of motion diversity.

### Dimensionality reduction of cardiac motion

Three dimensional time-resolved point clouds capturing cardiac motion are spatially and temporally correlated. Dimensionality reduction was performed to extract a low-dimensional abstract representation capturing the variance in motion traits over time. The entire pipeline from high-dimensional cardiac motion representation to low-dimensional interpretable signatures is described in Figure 3.

**Figure 3.**
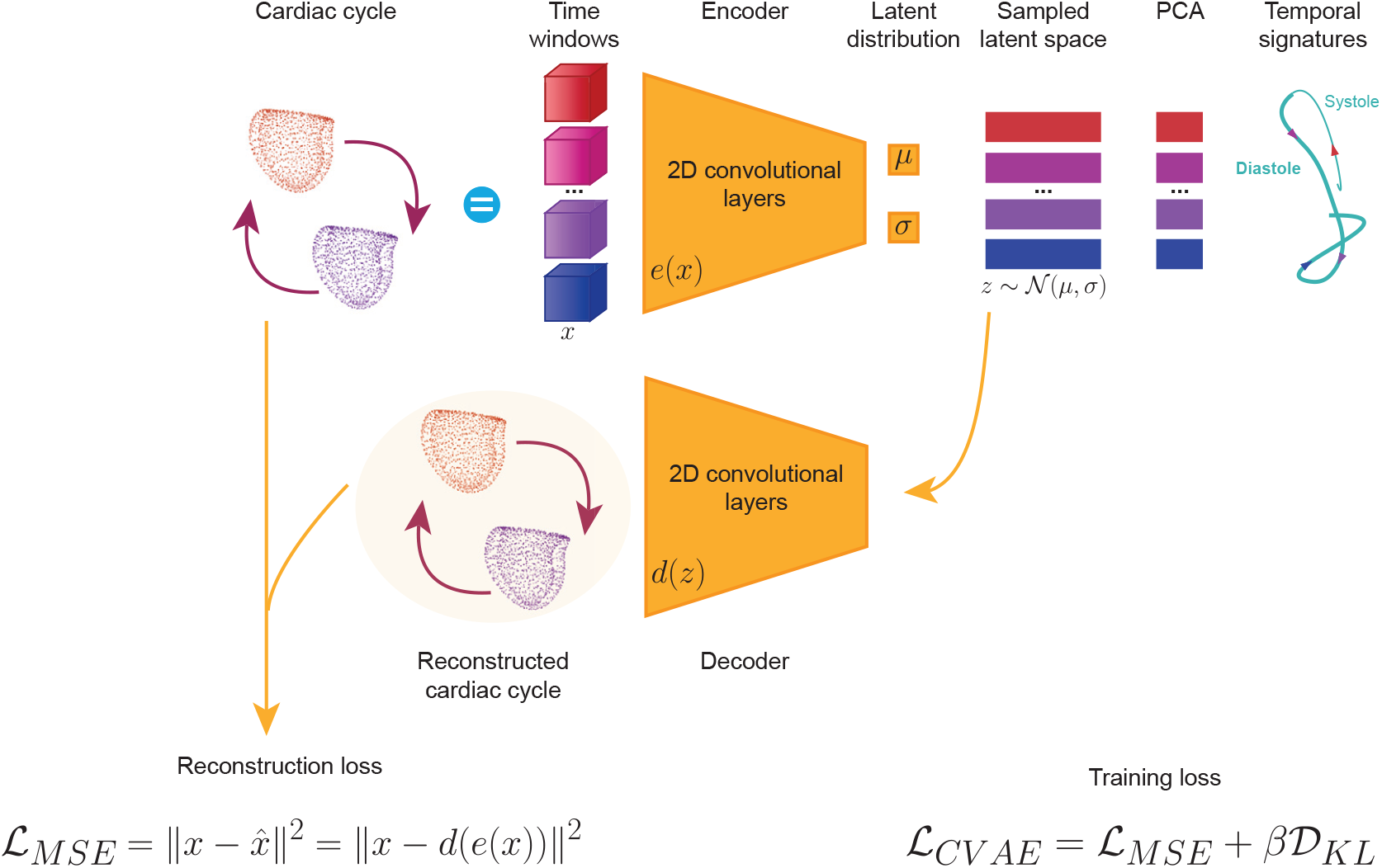
Convolutional variational autoencoder (CVAE) and temporal signatures. The cardiac cycle was fed in the model as a collection of time windows. The encoder and decoder were made of two-dimensional convolution layers. The encoder took as input the collection of windows and encoded a latent distribution of mean *μ* and standard deviation *σ*. The latent space *z* was sampled from that distribution and was fed back into the model through the decoder. The decoder aimed to reconstruct the input cardiac cycle. The reconstruction loss between input and model reconstruction was assessed using a mean squared error (MSE). The model was trained with the weighted sum of MSE and Kullback–Leibler (KL) divergence, which ensured the coherent continuity of the latent space. After training, the dimensionality of the latent space was further reduced with principal component analysis (PCA). Subsequent windows were concatenated and visualised in the reduced space in the form of temporal signatures describing the input cardiac cycle.

We used a variational autoencoder (VAE) with three convolutional layers in both the encoder and decoder. This network encodes a time window of cardiac motion in a latent vector and then decodes the vector into a reconstruction of that time window. Training the network to minimise the error between the reconstructed and input data ensures that the latent vector captures the complexity of the cardiac motion while reducing redundancy.

Post-training reconstruction loss, or relative mean absolute error, expressed in percentage of absolute range in each dimension was: 4.09±1.03% for X, 4.09±0.97% for Y, and 2.29±0.96% for Z coordinates. These values are comparable to observed errors in automated cardiac segmentation algorithms.^10^ Reconstruction was also evaluated post-training using a Chamfer distance (0.09±0.01) in normalised coordinates, which is comparable to the prediction error for advanced point cloud reconstruction algorithms.^11,12^

### Phenotypes associated with latent motion signatures

The latent space of the VAE is transformed using PCA. Reduced latent representations of consecutive time windows were concatenated into visually-interpretable temporal signatures or trajectories, characterising variations in complex cardiac motion traits. Such individual-level temporal signatures are low-dimensional uncorrelated abstractions of complex and highly-correlated moving point clouds.

The latent space was explored for associations with structural and functional features as well as circulating biomarkers (Supplementary Figure 1). The first principal component (PC1) was negatively correlated with body surface area, ventricular and atrial volumes, left ventricular mass, and wall thickness, and positively correlated with ejection fraction and high-density lipoprotein (HDL) cholesterol. PC2 was positively correlated with body surface area, ventricular volume, and mass. PC3 was positively correlated with age, and negatively correlated with ventricular and atrial volumes, left ventricular mass and wall thickness. PC4 was positively correlated with circumferential strain, ventricular volumes and mass and ejection fraction. These findings show how biophysical, structural and functional traits are embedded in a latent representation of heart motion.

A low dimensional motion signature was visualised using the first two components at each time point in the cardiac cycle. The position and morphology of the signature was represented across different z-scores of a given feature of interest in the population distribution (Figure 4). For instance, aging caused an in-plane translation of the signature towards lower values of PC1 and PC2, and a larger contour area. We observed opposing effects for hypertrophic cardiomyopathy (HCM) and dilated cardiomyopathy (DCM) polygenic risk scores in the latent space projection consistent with findings from studies assessing static morphological traits.^13,14^ Maximum wall thickness increased for low values of PC1, high values of PC2, and larger contour areas. The same trend was observed for systolic blood pressure along with greater differences in contour during systole (beginning of the signature), demonstrating that our model preserves temporal characteristics during dimensionality reduction of the cardiac cycle. HDL cholesterol increased for high values of PC1. Glycated hemoglobin (HbA1c), a marker of average blood sugar levels, rose with decreasing values of PC1 and PC2.

**Figure 4.**
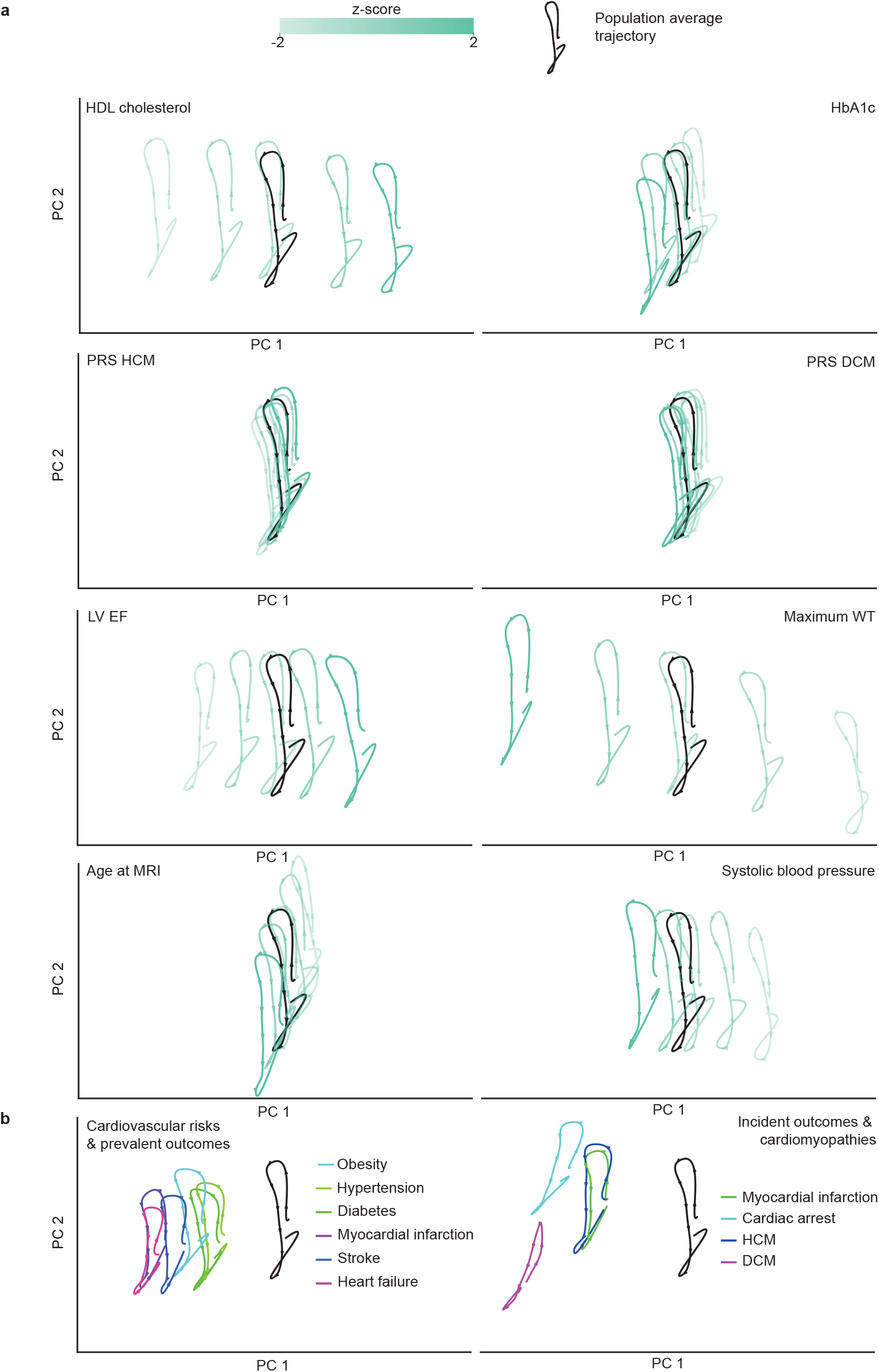
Signature distribution by feature z-score and signature average by binary outcome, on latent space position. **a**. Temporal signatures are plotted for relevant variables grouped by increasing z-scores. For each variable, individuals were stratified into five evenly spaced z-score bins within the range [−2, 2], and the mean latent signature was computed for each group. Feature variations shift signatures from the population average signature, in black on each plot. **b**. The average signature is plotted for each cardiovascular risk, prevalent and incident outcome. HDL, high-density lipoprotein; HbA1c, glycated hemoglobin; PRS, polygenic risk score; DCM, dilated cardiomyopathy; HCM, hypertrophic cardiomyopathy; LV, left ventricle; EF, ejection fraction; WT, wall thickness; MRI, magnetic resonance imaging.

Cardiovascular risk factors such as hypertension, obesity and diabetes were associated with low values of PC1. Adverse prevalent outcomes, including myocardial infarction, heart failure and stroke, resulted in a similar motion signature. Critical future outcomes, such as cardiac arrest, showed the lowest PC1 values and more extreme motion signatures. Incident HCM and incident myocardial infarction signatures were found to share the same in-plane position at low values of PC1. The average signature for future DCM was the furthest from the population average signature, with low values of both PC1 and PC2. These feature distributions showed how both the in-plane position and contour characteristics of the signature reflect the phenotype and genotype of participants, as well as prevalent and incident events.

### Clustering of motion phenogroups

Signatures were clustered on their PCA projection: first by contour characteristics and second by in-plane position as seen in Figure 5. K-means for time series data was used, and contour clustering identified two phenogroups, while in-plane position clustering revealed three, resulting in six clusters in total. Their relative in-plane positions are visualised in Figure 6a and their centred contours in Figure 6b. The most and least represented phenogroups represented 22.7% and 12.9% of the initial study population.

**Figure 5.**
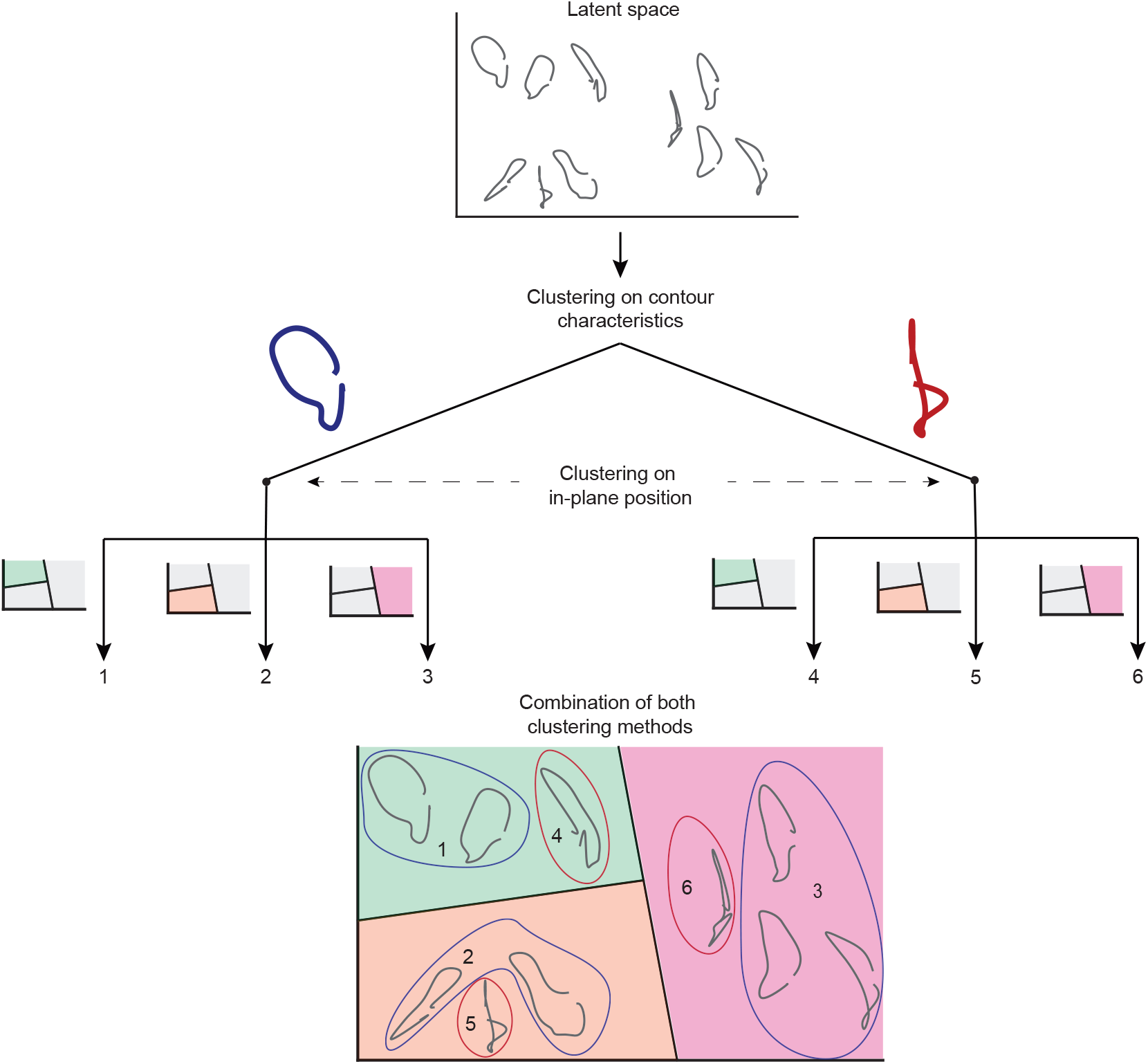
Combination of clustering on in-plane position and contour. Temporal signatures were clustered in two stages. A few individual examples are shown here to simplify the figure. First, the signatures were centred around the origin and clustered on their relative coordinates, capturing the contour of the trajectories, resulting in two clusters. Second, clustering was done on the original, uncentred coordinates of the temporal signatures, capturing their in-plane position, and revealing three clusters. The results of both methods were combined to give a dual clustering of the latent space, giving six clusters numbered from 1 to 6.

**Figure 6.**
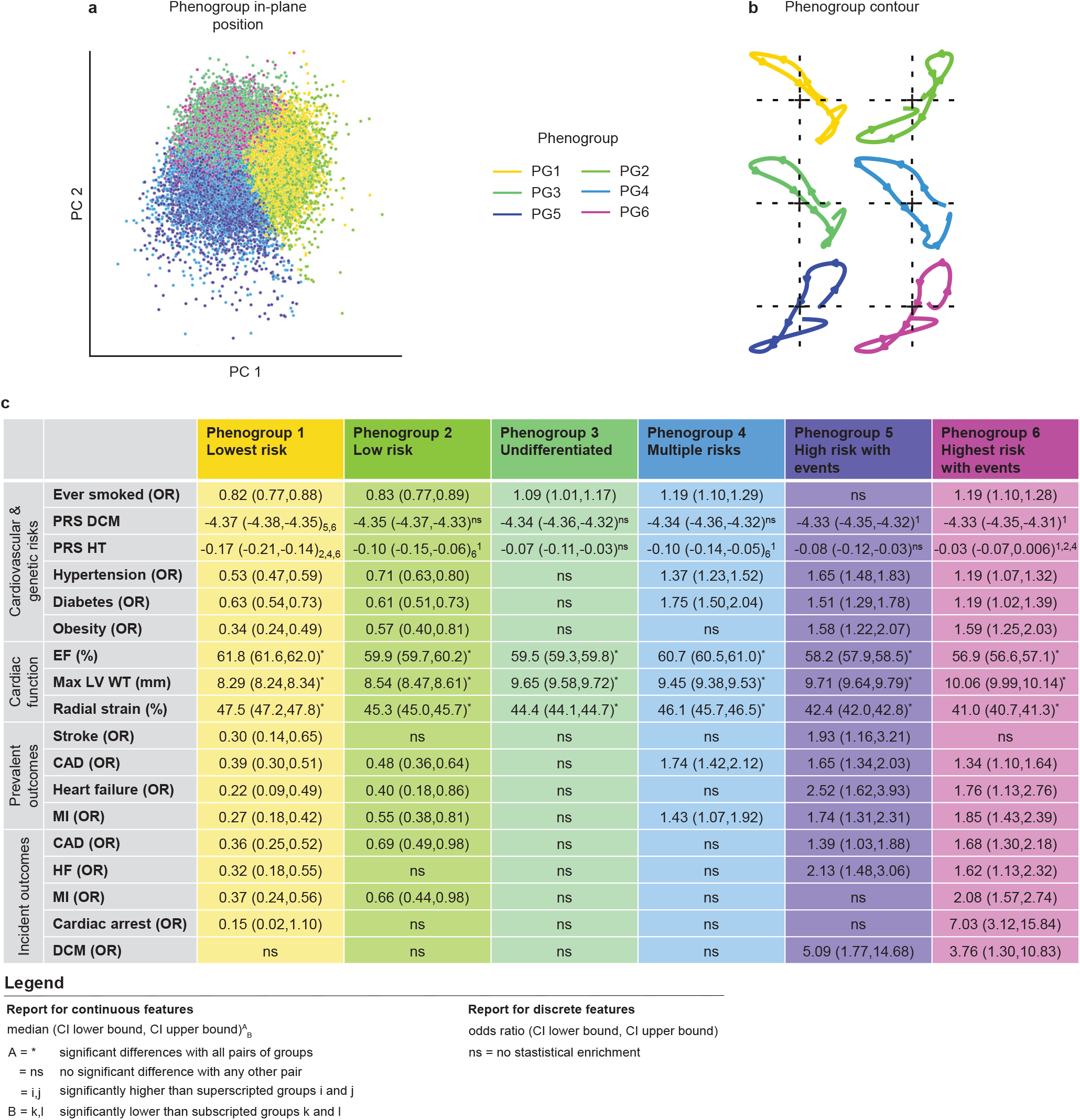
Clinical interpretation of motion-based phenogroups. **a**. Barycentre position of all individual trajectories, coloured by their assigned phenogroups, in the latent space of the two principal components. **b**. Average signatures for each phenogroup centred around the origin of the latent space. **c**. Summary of findings characterising the cardiovascular risks and disease status of the phenogroups. Participant characteristics and diagnostics were used to highlight the differences between groups. Findings are reported as the median confidence interval for continuous variables (Figure 7) and odds ratio and direction of association for discrete features (Figure 8) following statistical testing. A Kruskal-Wallis test followed by a post-hoc Dunn test was used for continuous variables, while a χ ^2^ test followed by a post-hoc Fischer test was performed for discrete variables. Multiple testing was accounted for by the Benjamini-Hochberg procedure, and the level of significance was *P* < 0.05. Sample size was n = 20,318. PRS DCM has been multiplied by 10^7^ for readability. PRS, polygenic risk score; DCM, dilated cardiomyopathy; HT, hypertension; EF, ejection fraction; LV, left ventricle; WT, wall thickness; mm, millimeters; CAD, coronary artery disease; MI, myocardial infarction; CI, confidence interval.

Individual-level features were tested for significant differences in the six phenogroups. Principal findings are summarised in Figure 7 for significantly different pairwise comparisons of continuous traits, and in Figure 8 for discrete variables and associated odds ratios (OR). Each odds ratio is derived from the contingency table of the χ ^2^ test performed to assess the enrichment of the observed discrete feature in one phenogroup against the rest of the population. A summary of findings for numerical features is found in Supplementary Figure 2. All the statistical test results are detailed in the Supplementary Data.

**Figure 7.**
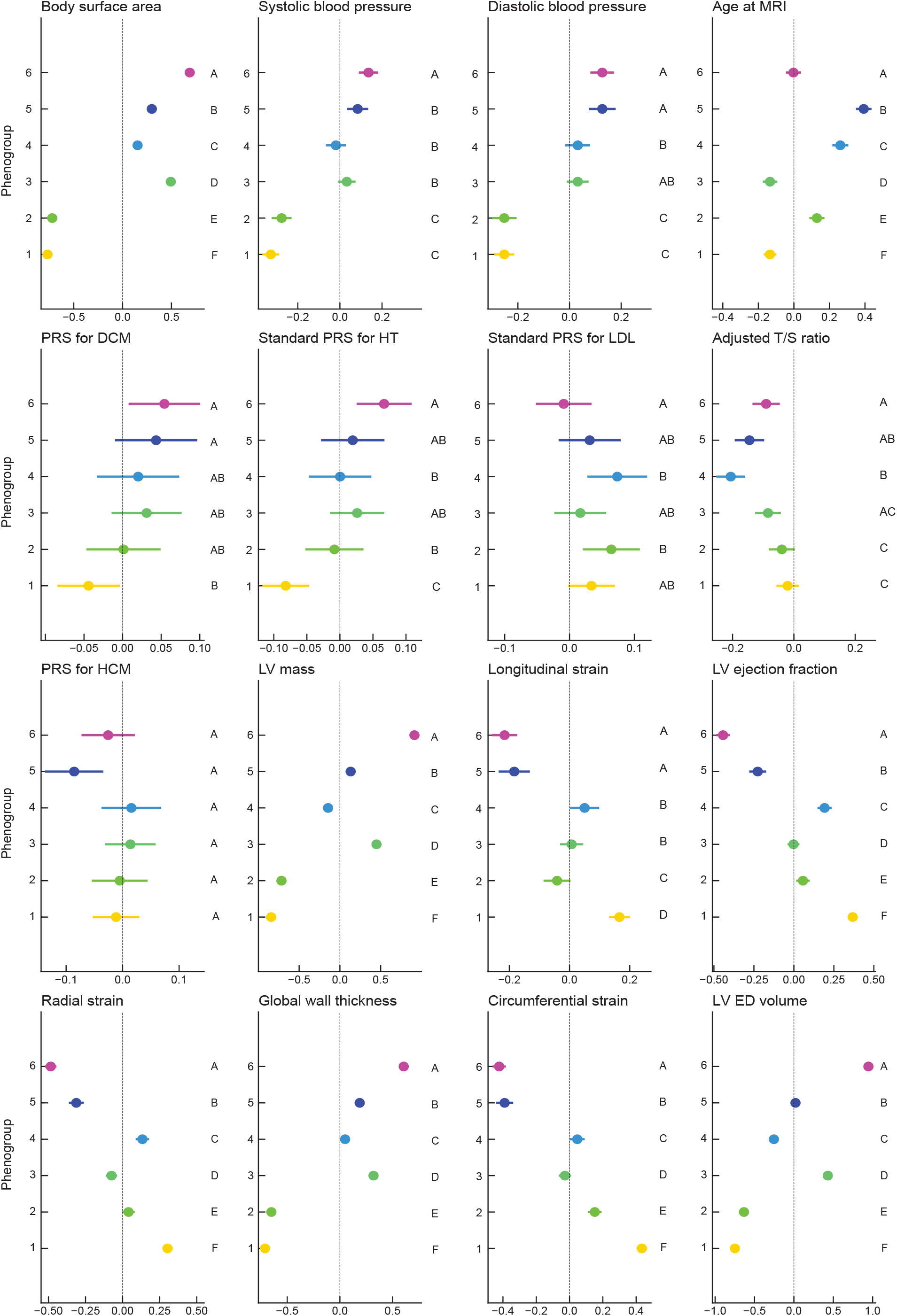
Numerical features significantly associated with motion-based phenogroups. Numerical features were tested for significant associations with the phenogroups. A Kruskal-Wallis test was performed, followed by a post-hoc Dunn test revealing pairs of phenogroups sharing significant differences in variable distribution. Benjamini-Hochberg correction for multiple testing was done, taking into consideration discrete features as well (see Supplementary Data for all tested features). The significance level was *P* < 0.05 and the sample size was n = 20,318. The median of the z-score and its confidence interval are displayed for each phenogroup. For a given feature, groups share a letter if there is no significant difference between them for that feature. MRI, magnetic resonance imaging; DCM, dilated cardiomyopathy; PRS, polygenic risk score; HT, hypertension; LDL, low-density lipoprotein; T/S, telomere to single copy gene; HCM, hypertrophic cardiomyopathy; LV, left ventricle; ED, end-diastolic.

**Figure 8.**
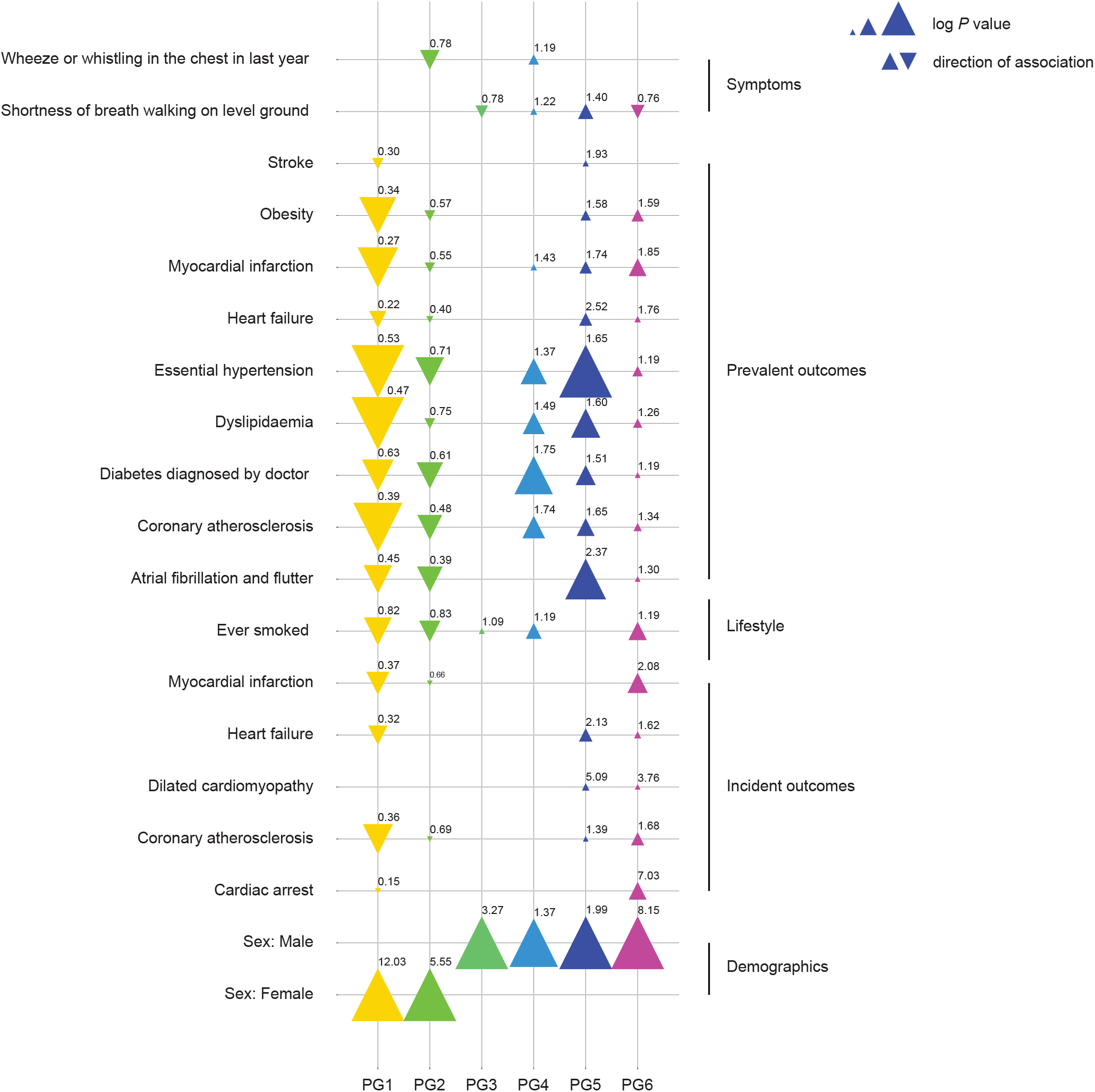
Discrete and binary features significantly associated with motion-based phenogroups. Discrete features were tested for significant associations with the phenogroups. A χ ^2^ test was used, followed by a post-hoc Fischer’s unit test which highlighted significant enrichment for discrete features. Benjamini-Hochberg correction for multiple testing was done, taking into consideration numerical features as well (refer to Supplementary Data for all tested features and detailed statistical results). The significance level was *P* < 0.05 and the sample size was n = 20,318. The size of the triangle reflects the logarithm of the *P* value obtained with each statistical test while its direction is of the association. The odds ratio is also displayed for each significant association.

Associations with risk factors and outcomes allowed us to construct an interpretative table of phenogroups from PG1 to PG6 ranked by increasing burden of cardiovascular risks (Figure 6c). Motion traits in PG1 were associated with reduced cardiovascular risk factors including hypertension (OR: 0.53; 95% confidence interval [CI]: 0.47, 0.59), diabetes (OR: 0.63; 95% CI: 0.54, 0.73), and obesity (OR: 0.34; 95% CI: 0.24, 0.49). It also exhibited the highest contractile function (radial strain (47.5%; 95% CI: 47.2, 47.8) and ejection fraction (61.8%; 95% CI: 61.6, 62)). In PG2 cardiometabolic risks were slightly higher (hypertension (OR: 0.71; 95% CI: 0.63, 0.80), diabetes (OR: 0.61; 95% CI: 0.51, 0.73), and obesity (OR: 0.57; 95% CI: 0.40, 0.81)). The least differentiated group for risk factors was PG3 with no significant associations. PG4 had the highest risk of diabetes (OR 1.75; 95% CI: 1.50, 2.04), and was also enriched for smoking (OR: 1.19; 95% CI: 1.10, 1.29) and hypertension (OR: 1.37; 95% CI: 1.48, 1.83). This group also had a significant risk for prevalent coronary artery disease (CAD) (OR: 1.74; 95% CI: 1.42, 2.12) and myocardial infarction (OR: 1.43; 95% CI: 1.07, 1.92).

PG5 had the highest risk for hypertension (OR: 1.56; 95% CI: 1.48, 1.83), and was also associated with diabetes (OR: 1.51; 95% CI: 1.29, 1.78) and obesity (OR: 1.58; 95% CI: 1.22, 2.07). This group also had lower ejection fraction (58.2%; 95% CI: [57.9, 58.5]) and radial strain (42.4; 95% CI: [42, 42.8]). PG5 had the most enrichment for stroke (OR: 1.93; 95% CI: 1.16, 3.21), prevalent (OR: 2.52; 95% CI: 1.62, 3.93) and incident (OR: 2.13; 95% CI: 1.48, 3.06) heart failure, and a future diagnosis of DCM (OR: 5.09; 95% CI: 1.77, 14.68).

Lastly, PG6 was linked to genetic risk factors including polygenic scores for hypertension and DCM. This group had the lowest ejection fraction (56.9%; 95% CI: [56.6, 57.1]) and radial strain (41; 95% CI: [40.7, 41.3]), as well as the highest maximum wall thickness (10.1 mm; 95% CI: [9.99,10.14]). PG6 was enriched for conventional risk factors, as well prevalent adverse cardiovascular outcomes: CAD (OR: 1.34; 95% CI: 1.10, 1.64), heart failure (OR: 1.76; 95% CI: 1.13, 2.76), and myocardial infarction (OR: 1.85; 95% CI: 1.43, 2.39). PG6 had the most associations with incident outcomes of all the phenogroups: CAD (OR: 1.68; 95% CI: 1.30, 2.18), heart failure (OR: 1.62; 95% CI: 1.13, 2.32), myocardial infarction (OR: 2.08; 95% CI: 1.57, 2.74), cardiac arrest (OR: 7.03; 95% CI: 3.12, 15.84) and a diagnosis of DCM (OR: 3.76; 95% CI: 1.30, 10.83).

### Method stability and benchmark models

A stability analysis was performed to assess the robustness of the clustering method. The analysis also revealed the optimal number of clusters as per the maximum average Rand index (ARI) score^15^, which was three for the in-plane position clustering (Supplementary Table 1) and two for the trajectory contour clustering (Supplementary Table 2). The ARI scores range from 0 to 1, with 1 being a perfect match between two clustered sets.

Our method was compared to two benchmark models. First, we swapped the input data of high-dimensional point clouds for 28 different CMR-derived tabular measures spanning volumes, mass and strain (Supplementary Table 3) to show that latent representations of motion contain more clinically-relevant information. Second, we changed our dimensionality reduction method to a simple linear PCA applied directly on the cardiac point clouds.

The comparison of our framework to the two benchmark models is summarised in Supplementary Table 4 with our framework having an ARI of 0.909. Using CMR-derived clinical measures as input, UMAP (uniform manifold approximation and projection)^16^ was applied to reduce the dataset dimension to 16, the same as our model bottleneck, followed by PCA. A K-means algorithm was used to cluster the resulting principal components. This resulted in a lower ARI of 0.82. Using simple PCA to reduce dimension, keeping everything else equal, gave an ARI of only 0.50 and 0.85 for clustering by in-plane position and contour, respectively, giving a less stable method when comparing with our scores of 0.914 and 0.904.

### Visualising phenogroup motion patterns

Phenogroup motion patterns were computed by comparing average point velocity of the ventricular surface to the population average (inferolateral: Figure 9b and anteroseptal: Supplementary Figure 3). In the higher risk groups we observed characteristic changes in patterns of cardiac motion. In PG4, where cardiometabolic risks were highest, we observed reduced velocities in the basal regions of the heart in both systole and diastole suggesting there are regionally localised changes. For PG5 reduced velocities were more globally distributed in the left ventricle but confined predominately to systole and early diastole. In contrast, for PG6 with the highest burden of poor outcomes, reduced function extended throughout diastole as well. The lowest risk phenogroups show comparatively faster systolic velocities especially in the basal ventricle as well as more rapid diastolic relaxation. These findings show how spatio-temporal patterns of motion vary between the discovered clusters.

**Figure 9.**
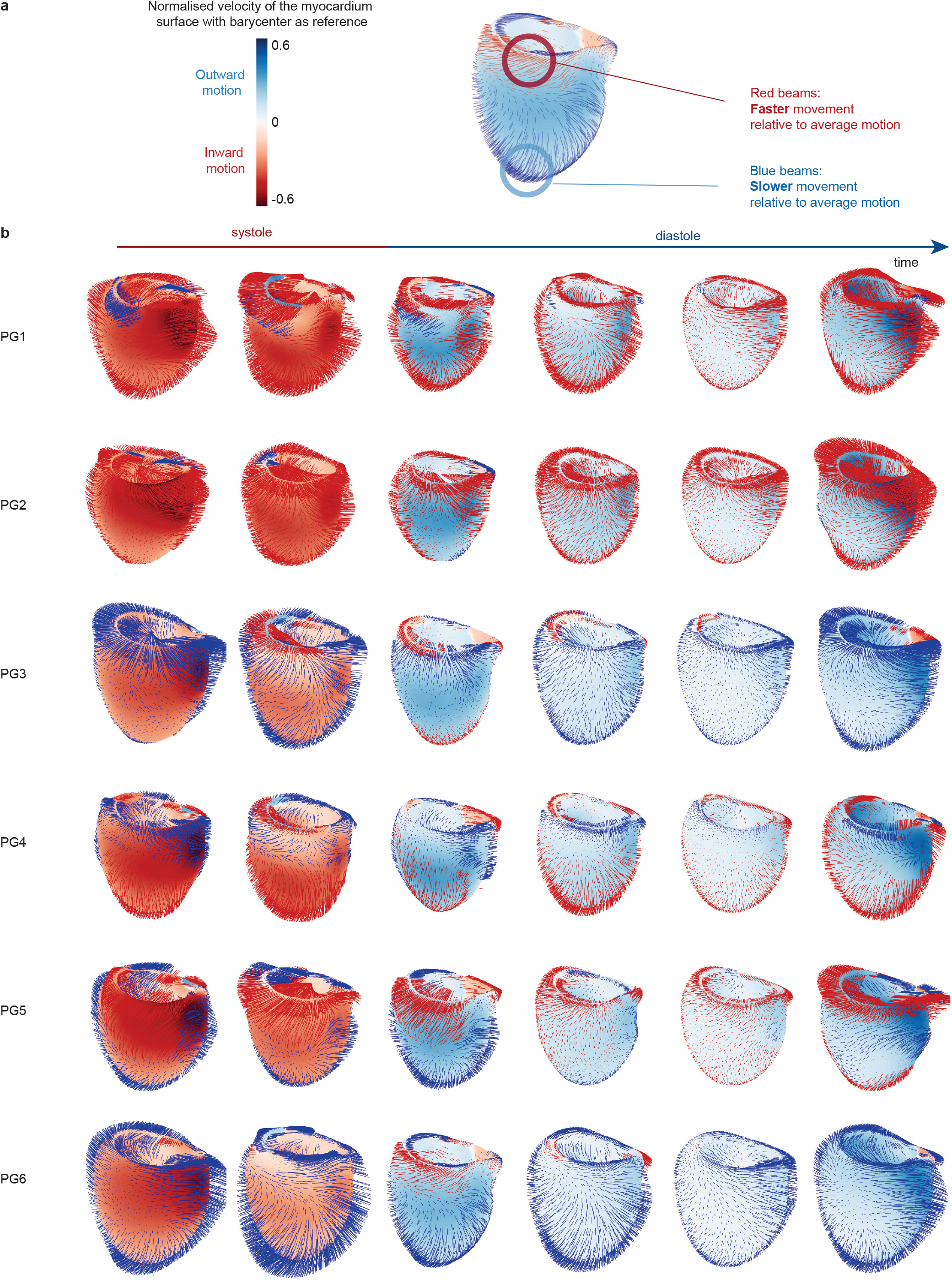
Left ventricle motion velocity by phenogroup - Inferolateral view. **a**. Zoomed in examples of the left ventricle during systole and diastole. The velocity of the ventricle wall is displayed on the surface, relative to its barycentre. The velocity is positive during relaxation, or diastole when the ventricle is filling with blood, while it is negative during contraction, or systole when ventricular ejection occurs. Red beams indicate areas of faster movement relative to the population average motion, while blue beams show areas of relative slower movement. **b**. The left ventricle motion was compared to the population’s average motion for each phenogroup.. The six frames represent one entire cardiac cycle from systole (contraction of the ventricle) to diastole (relaxation of the ventricle). The velocity of the ventricle was calculated as the local gradient of change between two consecutive frames, averaged out over multiple frames to reduce the number of displayed frames, and normalised for the size of the ventricle.

## Discussion

The heart is a highly complex electromechanical organ system but conventional assessment of dynamic contraction relies on pre-defined volumetric, deformational and flow parameters that may not provide efficient evaluation of early dysfunction or advancing disease. Our study shows there is latent information in point cloud data representing patterns of cardiac motion that predict future events, capture the effect of risk factors on function, and provide a new classification of motion-based traits in an adult population. This approach also produces human-interpretable signatures to assist in understanding how function deviates across multiple dimensions of health.

Ejection fraction is an established marker of global ventricular function which is representative of volumetric approaches for assessing cardiac status and defining a diagnosis of heart failure.^3,17^ It is also poorly predictive of symptoms or treatment response with various thresholds used to define putative underlying mechanisms.^18,19^ Myocardial strain is a more sensitive marker of contractile function, can be measured globally or regionally, and is independently predictive of outcomes.^20^ However, strain is purely a measure of myocardial shortening or thickening,^21^ and offers only a narrow assessment of cardiac mechanics. Statistical shape modelling of the heart hints at the potential for even static whole-heart features to encode latent information on genetic regulation.^7,22,23^ Here we use a strategy of creating a dynamic three dimensional point cloud, densely sampling the left ventricle, to capture a rich feature space of motion throughout the whole cardiac cycle. This concept applied to patients with heart failure is strongly predictive of outcomes,^1^ but has not previously been used in large populations or for discovery of disease mechanisms. Clustering of conventional image-derived phenotypes has found mechanistically-distinct groups in patients with DCM,^24^ and congenital heart disease for instance.^25^ Clustering of static three-dimensional shape data can also reveal mechanistic and prognostic patterns in HCM,^26^ but there is limited knowledge of undertaking this on moving point clouds. To achieve this we created a latent space of cardiac motion using a task-specific VAE framework and visualised this as a low dimensional signature that could be used for stable clustering.

We found that a low dimensional representation of whole-heart motion captured associations with circulating biomarkers for diabetes and coronary artery disease. Established diabetes has been causally linked to diastolic dysfunction,^27^ suggesting that our models are sensitive to changes in myocardial stiffness without relying on predefined measures of relaxation. We also assessed polygenic scores that estimate predisposition to cardiomyopathies that are defined by static phenotypes, finding that they also have an effect on cardiac motion. The association between common genetic variants and heart motion in the adult population is not well understood but abnormal contractility, diastolic dysfunction and dyssynchrony are recognised in established disease.^28^ Conventional cardiovascular risk factors such as obesity and hypertension were associated with motion traits and we found enrichment for future cardiac events in the latent space. We also observed sex-related differences in motion traits highlighting the potential for three-dimensional analysis in revealing sex-specific functional characteristics that may underlie variance in disease penetrance.^29^ Taken together, this shows the potential for unsupervised motion analysis to discover features that relate to a range of environmental and genetic risk factors which are also prognostically important.

Clustering identified six phenogroups with varying enrichment for cardiovascular risk factors and outcomes. The lowest risk groups (PG1 and PG2) had motion traits associated with the least risk for obesity, diabetes and hypertension, as well as both prevalent and incident outcomes. This suggests that healthy individuals with a low risk of adverse outcomes can be identified through motion analysis, and that the approach is not limited to those with established or sub-clinical disease. The cluster most strongly enriched for cardiometabolic disease was PG4. Diabetic cardiomyopathy is related to hypertrophy and fibrosis, early-onset diastolic dysfunction and late-onset systolic dysfunction.^30^ Here we show even in a broad population of community-living adults that motion traits can be associated with metabolic risk. The last two clusters (PG5 and PG6) had the strongest enrichment for both prevalent and incident endpoints, as well highest polygenic risks. PG6 was the only cluster marked by higher incident myocardial infarction and cardiac arrest, suggesting that motion traits are predictive of fatal outcomes. Conventional parameters including ejection fraction and radial strain were lowest in PG5 and PG6 but we found that even a broad range of CMR parameters did not result in stable clustering.

Our convolutional VAE generates temporal embeddings from time windows of cardiac motion data, and avoids flattening temporal data before passing it to the model. Our model also provides interpretability by preserving temporal characteristics of the data in its output. This methodology could be extended to longitudinal imaging datasets to reveal temporal trajectories of disease progression, further improving risk modelling and targeted clinical interventions. There are some limitations to the input data. While the CMR contained 50 temporal frames it does not have the temporal resolution of echocardiography. It is also not as widely available for inference as echocardiography. The population being studied also has limitations including under-representation of older age groups and persons living in less socioeconomically deprived areas. The UK Biobank population is also predominantly European and further work is required in people of diverse ancestries and social groups. We performed a robust analysis of model stability and comparison to a benchmark of conventional parameters but there is no equivalent data available at scale for external validation.

In conclusion, we show how time-resolved three dimensional representations of cardiac motion contain rich information on cardiovascular risks, disease mechanisms and outcomes while providing human interpretable signatures.

## Methods

### UK Biobank participants

UK Biobank is a large cohort study on half a million individuals in the UK aged 40-69 at recruitment between 2006 and 2010.^31^ All participants provided written informed consent for the study, which was also approved by the National Research Ethics Service (11/NW/0382). Our study was conducted under terms of access approval number 28807 and 40616. A collection of a wide range of phenotypic and genotypic attributes, risk factors and physical measures is available. They were obtained by touch screen questionnaires, interview, biophysical measurement, hospital episode statistics, and primary care data. CMR images were also acquired for a subset of participants. Details of how each phenotype was acquired are available on the UK Biobank Showcase (http://biobank.ctsu.ox.ac.uk/crystal/).

Cardiovascular diseases and outcomes were derived from groupings of ICD-10 codes as described in Supplementary Table 5. Outcomes were classified as prevalent or incident if they happened before or after the imaging visit. Polygenic risk scores (PRS) for HCM and DCM were derived from genetic sequencing data identifying rare pathogenic and likely pathogenic variants in the UK Biobank population and other cohorts.^32–34^ All the other mentioned PRS were accessed through the UK Biobank PRS Release.^35^

### Cardiac imaging segmentation and strain analysis

CMR imaging was performed on participants to capture two-dimensional retrospectively-gated cine imaging on a 1.5T magnet (Siemens Healthineers, Erlangen, Germany).^36^ Left ventricular short-axis plane cine images from base to apex were acquired as well as long-axis cine images in the two and four-chamber views. Cines comprised 50 cardiac frames with a typical temporal resolution of 31 ms. Ascending and descending thoracic aorta images were also acquired with transverse cine imaging.

The two-dimensional cine images in the short and long-axis were segmented using fully convolutional networks, with segmentation quality equivalent to expert human readers.^9^ Derived measurements of the left and right ventricles were determined from the two-dimensional images: end-diastolic and end-systolic volumes, stroke volume, and ejection fraction. A density of 1.05 g.ml^-1^ for the left ventricular myocardium was used to derive its mass from its volume. With *A*_2*Ch*_ and *A*_4*Ch*_ as the atrial areas on the two and four-chamber cine views respectively, and *L* as the averaged longitudinal diameter across two views, the atrial volumes were derived as per the following formula on biplane area-length: 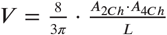 CMR-derived measurements were indexed to body surface area (BSA) calculated with the Du Bois formula: BSA = 0.007184 * Height^0.725^ * Weight^0.425^ with height in cm and weight in kg. The wall thickness of the left ventricular myocardium was measured as the distance between the segmented epicardium and endocardium at end-diastole.

Circumferential and radial strains were calculated with cine short-axis images as per the formula 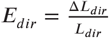 with *dir* the circumferential or radial direction, *L*_*dir*_ the length of a line segment in this direction and Δ*L* _*dir*_ the change of length over time. Longitudinal strain was derived from long-axis four-chamber motion tracking.

A spatio-temporal neural network was used to segment the aorta cine images.^37^ Maximum and minimum cross-sectional areas were derived and aorta distensibility was calculated with central blood pressure obtained using peripheral pulse-wave analysis (Vicorder, Wuerzburg, Germany).^38^

### Three dimensional motion capture

Motion was represented in a three-dimensional biventricular model by fitting a shape-refined mesh to 2D segmentations and aligning these to a reference atlas. This atlas was defined from over 1000 healthy adults which enforced spatial and temporal consistency.^39^ The processing pipeline consists of three main steps, as illustrated in Figure 2.

### Segmentation

A multi-task deep learning model was used to simultaneously generate anatomical labels and key landmarks on the CMR data.^40^ To enhance anatomical accuracy, prior shape information was incorporated through label propagation from high-resolution anatomical atlases. Here a three dimensional high-resolution atlas was non-rigidly registered to each participant’s end diastolic frame to obtain a sub-voxel-smooth segmentation following atlas selection based on landmark localisation and shape similarity.

### Mesh fitting

After segmentation, each participant’s data was aligned to a reference atlas space using a series of image registration steps, including affine and non-rigid B-spline transformations. This process ensured spatial consistency across the cardiac cycle and preserved anatomical correspondence of mesh vertices over time. A subject-specific end-diastolic mesh was generated by non-rigidly fitting the atlas mesh to the segmentation surface reconstructed using a marching-cubes algorithm.

### Three dimensional time-resolved motion estimation

Cardiac motion was estimated using a “four dimensional” spatio-temporal free-form deformation applied to the cine images with sparsity regularisation.^39^ Displacement fields were computed between successive time frames in both forward and backward directions to reduce alignment errors. These motion fields were applied to the subject-specific mesh to generate a sequence of 50 deformed surface meshes each corresponding to a distinct time point in the cardiac cycle. Temporal normalisation was performed to ensure consistency across frames resulting in meshes with identical vertex counts that maintain anatomical correspondence both temporally and spatially across participants.

### Data pre-processing

Point clouds were extracted from the cardiac meshes as unstructured collections of points. The point clouds described the movement of the left ventricle myocardium during one periodic cardiac cycle. Point clouds were down-sampled by a 97% factor using quadratic clustering in Python using the vtk package, from 40,616 to 1406 points. This does not affect any subsequent analysis as demonstrated in a previous study using cardiac motion data for prediction and risk-stratification.^1^ Point clouds were then transformed to fit in a unit sphere: they were centred around the origin by subtracting their centroid and range-normalised by dividing the coordinates by the distance of the most distant point to the centroid across the whole dataset. This pre-processing allows the preservation of intra-population size variability while translating all data points between -1 and 1, which is needed before feeding it into our neural network. Windows of cardiac motion were constructed along the cardiac cycle using a sliding window of 16 frames, a stride of 4 frames and padding at the end to ensure periodicity (Supplementary Figure 4a). These parameters were chosen to capture an accurate representation of the cardiac cycle in each window while limiting the use of computational power. Consecutive windows overlap each other to ensure the continuity of the cardiac cycle. This resulted in 13 windows for each cardiac cycle as per the ceiling value of the number of total frames divided by the stride. The 1406 point coordinates were down-sampled to 1024 to match the architecture of the autoencoder.

### Convolutional variational autoencoder

The input *x* of the model is a 3-channel tensor of size 3 × 16 × 1024, where the three channels represent the X, Y and Z coordinates, 16 is the number of time points in a window, and 1024 is the number of points in each point cloud (Supplementary Figure 4b).

A convolutional variational autoencoder (CVAE) was implemented using PyTorch to encode these spatio-temporal matrices into a compact latent representation. The encoder and decoder were made of successive 2D convolutional and transposed convolutional layers, respectively. 2D convolutions were employed due to the inherent two-dimensional structure of the reformatted data, with time points along one axis and spatial points along the other, enabling the model to learn local spatial and temporal features hierarchically. The encoder used four 2D convolutional layers with kernel size 4, stride 2 and zero padding. The number of feature channels was progressively doubled after each convolution, starting from 8 channels in the first layer. This hierarchical structure allowed the model to capture increasingly abstract features. The architecture of the model is described in detail in Supplementary Figure 5. Following the convolutional layers, an adaptive average pooling layer reduced each feature map to a single value, irrespective of its input size. This operation produced a fixed-size feature vector that is invariant to small changes in input dimensions, facilitating processing by subsequent fully connected layers. Adaptive pooling is suitable in our case as it standardises feature map sizes before flattening, simplifying the subsequent latent space parametrisation. The pooled feature vector was then passed through two parallel fully connected layers to produce the latent parameters: the mean vector *μ* and the standard deviation vector *σ* of a multivariate Gaussian distribution. The latent vector *z* was then sampled using the reparametrisation trick:

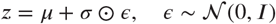

where *⊙* denotes element-wise multiplication. This technique allows gradient to be propagated through the stochastic sampling operation by expressing the random variable *z* as a deterministic function of *μ, σ* and a noise term *ϵ*.^41^ The decoder mirrored the architecture of the encoder, beginning with a fully connected layer that projected the latent vector back to a higher-dimensional feature space. The output was reshaped and successively upsampled using four 2D transposed convolutional layers with the same kernel size, stride, and padding settings as the encoder, progressively halving the number of channels while restoring the original matrix dimensions. This design choice allowed the model to reconstruct the spatial configuration of the cardiac geometry while leveraging the hierarchical features captured by the encoder.

The CVAE was optimised using a loss function combining a reconstruction loss and a Kullback-Leibler (KL) divergence term as per the *β*-VAE framework^42,43^:

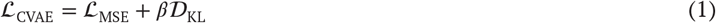

The reconstruction loss, ℒ_*MSE*_, was computed as the mean squared error (MSE) between the input point cloud *x* and 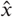: 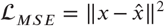. We selected MSE as it directly penalised large reconstruction errors and is suited for continuous numerical data such as point coordinates, and corresponds to assuming a Gaussian likelihood on the reconstruction error, which is consistent with the probabilistic framework of variational autoencoders. The KL divergence term, 𝒟_*KL*_, measures the divergence between the approximate posterior parametrised by *μ* and *σ*, and a multivariate standard normal prior:

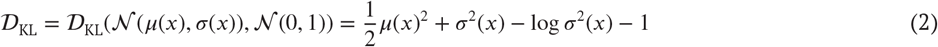

This regularisation encourages the latent space to conform to a known and structured distribution. This promoted generalisability to avoid overfitting and enabled interpolation between latent representations.^44^ The weighting parameter *β* controlled the trade-off between reconstruction accuracy and latent space regularisation. A higher *β* enforces stronger latent space regularisation, potentially at the cost of reconstruction quality, while a lower *β* prioritises reconstruction. We selected *β* = 0.001 after testing different values from 0.1 to 10^−5^. Finally, the model was trained on an NVIDIA RTX A6000 with an Adam optimiser, with an 80-20 split between training and validation, using a batch size of 256 and a learning rate of 0.001.

The quality of reconstruction of the cardiac point cloud time windows was quantitatively assessed post-training using two complementary metrics: the relative mean absolute error (MAE) and the Chamfer distance. For each time window, the absolute error between the input point cloud *x* and its reconstruction 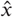 was computed as:

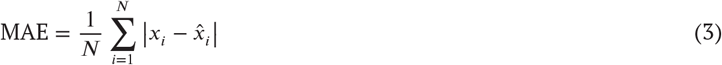

where *N* is the total number of points in the point cloud, | · | is the Euclidean distance, and 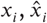 represent the 3D coordinates of the *i*^*th*^ point in the input and reconstruction, respectively. To enable interpretable error comparisons across participants with potentially different cardiac sizes and anatomical ranges, the MAE was normalised by the absolute range of the data in each dimension specific to each individual participant. This was computed as:

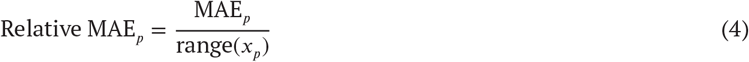

where range(*x*_*p*_) represents the range of in the input point cloud for participant *p*, computed as the difference between the maximum and minimum values along each spatial dimension, aggregated across all time windows for that participant. This normalisation gave a relative error expressed as a percentage of the anatomical data range of each participant, preserving inter-individual variability while ensuring interpretability and direct comparison across the cohort of the reconstruction error.

Additionally, we computed the Chamfer distance to further characterise the geometric similarity between the input and reconstruction as this metric is widely used for measuring distances between two point sets. Denoting the input point cloud *P* and the reconstructed point cloud *R*, the Chamfer distance is defined as^45^:

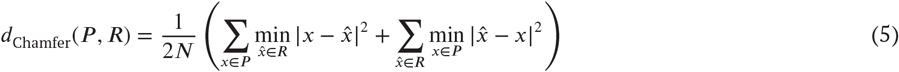

where |·| is the Euclidean distance, and N is the constant number of points in the point clouds. The metric computed the average squared nearest-neighbour distance between points in one cloud to the other, in both directions. The Chamfer distance captures both global and local discrepancies in the point cloud structure, independent of point ordering. This makes it particularly suitable for evaluating 3D anatomical reconstructions where point correspondence is implicit but not explicitly indexed. Unlike purely point-wise error metrics, Chamfer distance evaluated the overall spatial distribution of the reconstructed geometries. By combining participant-relative MAE and Chamfer distance, we obtain a complementary quantitative assessment of the performance of our model: the relative MAE is sensitive to normalised point-wise errors within the anatomical context while the Chamfer distance captures the structural geometric fidelity of the reconstructions.

### Latent space generation

The resulting latent space was normalised and PCA was performed to remove any correlation between the latent vectors. This facilitated further dimensionality reduction needed for visual representation and clustering of low-dimensional time series, as well as improved the interpretability of the latent features characterising the cardiac cycle. First, ten principal components were kept while preserving 99% of the initial variance. Consecutive latent representations were concatenated for each patient to form an abstract trajectory of embeddings, called temporal signatures (Figure 3). The signatures can be visually inspected in the reduced space of the first and second principal components accounting for 82% of the variance. As the cardiac cycle frames are ordered from the beginning of systole to the end of diastole in our dataset, signatures are similarly moving from systole to diastole. Only the first four principal components were kept for further analysis steps as they represented 98% of the variance, which was deemed an appropriate cut-off for representing the data while considering computational resources.

### Phenotype associations with temporal signatures

Temporal signatures are generated in an unsupervised way and are only abstract representations of the high-dimensional point clouds. Relevant participant features were tested for correlation with the latent principal components to explain the latent space variance. Data from demographics, body and clinical measurements, biomarkers and MRI-derived measures were accounted for. A subset of relevant features were also visually inspected with signature distribution stratified by z-score in the latent space of the first and second principal components: age, systolic blood pressure, maximum wall thickness, left ventricle ejection fraction, HDL cholesterol, HbA1c and PRS for HCM and DCM. For each feature, z-scores were computed as 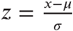 where x is the feature value of the individual, *μ* is the mean, and *σ* is the standard deviation of that feature across the population. The trajectories were grouped into five evenly spaced z-score bins within the range [−2, 2], and the average latent signature was computed within each bin. The average signature for selected risk factors and outcomes were also displayed in the latent space: hypertension, obesity, diabetes, stroke, heart failure, myocardial infarction, cardiac arrest, HCM and DCM.

### Unsupervised clustering of signatures

The latent representations capture the dynamic evolution of cardiac point clouds over time. These temporal signatures contain information on both the in-plane position and contour geometry of the trajectories in latent space. Importantly, as the temporal signatures are sequences of low-dimensional latent coordinates, they can be assimilated to multivariate time series, enabling the use of special time series clustering techniques.

Clustering of the temporal signatures was performed using K-means applied to time series data, implemented with the tsmeans^46^ Python library. This algorithm is an extension of the classic K-means clustering method, adapted for temporal data by replacing the standard Euclidean distance with a time series-specific distance measure of dynamic time warping (DTW). DTW is particularly well-suited for time series data as it accounts for potential temporal misalignments between sequences by finding an optimal non-linear alignment.^47,48^

We denote our set of *n* time series *s*_1_, *s*_2_, …, *s*_*n*_ for each participant, where each _*i*_ ∈ ℝ^*T* ×*d*^ represents a temporal signature of length *T* = 13 in *d* = 4-dimensional latent space (the first four principal components). The objective is to partition the time series into *K* clusters *C*_1_, *C*_2_, …, *C*_*K*_ to minimise the within-cluster sum of DTW distances:

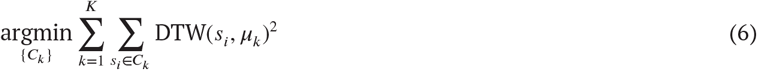

where *μ*_*k*_ ∈ ℝ^13×4^ is the centroid of cluster *C*_*k*_. DTW calculates the similarity between two time series by warping their time axis to achieve the best possible alignment. The metric is computed by finding the warping path 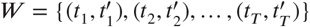 which minimises the distance:

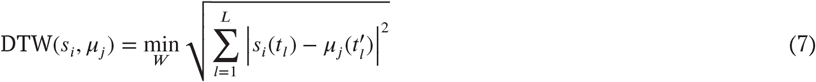

Signatures were clustered based on two properties: in-plane position and contour. First, temporal signatures were centred around the origin by subtracting their mean. This operation removed information about their absolute position in the latent space, isolating the relative contour of the trajectory. Clustering was then performed on these centred time series to group signatures with similar temporal evolution patterns, irrespective of their absolute position. After that, clustering was performed on the original, uncentred temporal signatures, which preserved information about the absolute in-plane position of the trajectories in latent space. This allowed grouping based on spatial positioning. The rationale behind this dual clustering approach lies in the hypothesis that both the contour geometry of the signature and its position in latent space convey clinically or physiologically distinct information. By combining the results of both methods, the final phenogroups integrate both geometrical and structural characteristics of the trajectories (Figure 5).

Stability analysis was performed to assess the robustness and reproducibility of the clustering method. The initial dataset was partitioned into five equally-sized, non-overlapping sub-partitions. The clustering algorithm was independently performed on each of these five sub-partitions 𝒯^(1)^, 𝒯^(2)^, …, 𝒯^(5)^. The cluster labels obtained from each sub-partition, noted 𝒮^(*m*)^, *m* ranging from 1 to 5, were then compared to the cluster labels obtained from clustering the full dataset, noted 𝒮^ref^, treated as the reference clustering. The agreement between clustering solutions was evaluated with the adjusted Rand index (ARI) score.^15,49^ The ARI score measures the similarity between two clustering solutions by considering all possible pairs of data points and evaluating whether each pair is consistently assigned to the same cluster or to different clusters in both the reference clustering and the sub-partition clustering. For each sub-partition, we can denote *a* the number of pairs of temporal signatures that are assigned to the same cluster in both 𝒮^(*m*)^ and 𝒮^*ref*^ and *b* the number of pairs assigned to different clusters in both methods. The total number of possible pairs of temporal signatures in a sub-partition of size *n* is:

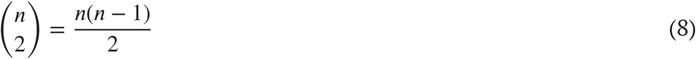

The Rand index (RI) is then calculated as:

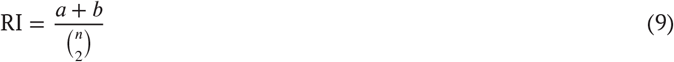

which gives the proportion of pairs on which the two clustering solutions agree. To adjust for random similarity agreement, the adjusted Rand index corrects the initial metric by subtracting the expected agreement between random clusterings and normalising by the maximum possible agreement:

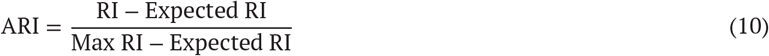

where the expected RI is the expected value for random clusterings, and the max RI is the maximum possible agreement. ARI ranges from -1 for a complete disagreement to 1 for a perfect agreement, with 0 indicating agreement expected by random chance. For each sub-partition, the ARI was computed by comparing 𝒮 ^(*m*)^ and 𝒮 ^*ref*^. The final stability score for both clustering on in-plane position and contour characteristics was obtained by averaging the ARI scores across the five sub-partitions:

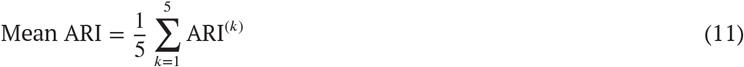

This analysis was repeated for different numbers of clusters *K* in the range of 2 ≤ *K* ≤ 5. This also allowed us to find the number of optimal clusters K for the K-means clustering algorithm applied to time series to ensure the maximum stability of the dual clustering method as per the ARI scores.

### Statistical testing for enrichment

Associations between phenogroups and a wide range of individual participant features were statistically tested. Features included demographics, lifestyle, risk factors, body and clinical measurements, MRI-derived measures, biomarkers, PRS, symptoms, outcomes and diagnosis. Longitudinal, radial and circumferential strains were considered as absolute values to simplify their interpretations. The full list of features can be found in the Supplementary Data. A Kruskal-Wallis test was used on continuous features to test for significant differences between the phenogroups. This was followed by a post-hoc Dunn test to find pairs of phenogroups with significant differences in feature distribution. A χ ^2^ test was used on binary and categorical features, followed by a post-hoc Fisher’s test to find significant associations of phenogroups with specific values of a feature. The effect size of each significant association was calculated as the odds ratio derived from the contingency table of the χ ^2^ test. The odds ratio measures the association of the outcome to the exposure of one phenogroup against the rest of the population. All resulting *P* values were adjusted for multiple comparisons under the Benjamini-Hochberg procedure, first for Kruskal-Wallis and χ ^2^ tests together, and then for post-hoc Dunn and Fischer tests together. Significant associations were reported for *P* value < 0.05. The Python packages used for the Kruskal-Wallis test, the post-hoc Dunn test and the Benjamini-Hochberg correction were respectively scipy, scikit-posthocs, and statsmodels. Additionally, a mass univariate group analysis was performed to compare geometry differences of temporal signatures between phenogroups. More details can be found in Supplementary Figure 6.

### Modelling motion in phenogroups

The velocity gradient of the left ventricle myocardium surface between consecutive point cloud frames was computed for the average cardiac motion in phenogroup. The average velocity of each phenogroup was normalised by dividing it by the bounding box size of the average heart for that group. Velocity differences were computed between each phenogroup and the population average cardiac motion. These variations were visually inspected across the cardiac cycle to identify unique motion patterns. The velocity fields were projected on the surface of the average motion by phenogroup to visualise outward and inward movements of the myocardium surface. Fields of beams were added onto the surface where the velocity of the myocardium was slower, for blue beams, or faster for read beams, than the population average velocity (Figure 9a). These four-dimensional models of motion differences helped identify distinctive motion patterns in each phenogroup.

## Funding

The study was supported by the Medical Research Council (MC_UP_1605/13); the Engineering & Physical Sciences Research Council (EP/W01842X/1); the British Heart Foundation (BBC/F/21/220106, RG/19/6/34387, RE/24/130023, CH/P/23/80008, FS/IPBSRF/22/27059, NH/F/23/70013); and the National Institute for Health Research (NIHR) Imperial College Biomedical Research Centre. For the purpose of open access, the authors have applied a Creative Commons Attribution (CC BY) licence to any author accepted manuscript version arising.

## Author contributions

Conceptualisation: P-R.S. and D.P.O.; Methodology: P-R.S., S.K., J.Z.; Software: P-R.S.; Validation: P-R.S.; Formal analysis: P-R.S.; Investigation: P-R.S.; Resources: D.P.O., P.I., K.A.M., S.L.Z., W.B.,; data curation: P-R.S., K.A.M., S.L.Z.; Writing - original draft: P-R.S. and D.P.O.; Writing - review & editing: all authors; Visualisation: P-R.S., D.P.O.; Supervision: J.S.W. and D.P.O.; Project administration: D.P.O.; Funding acquisition: D.P.O.

## Data availability

The analysis code repository is available on GitHub at: https://github.com/ImperialCollegeLondon/cardiac_motion_signatures/.

## Notes

### Competing Interest Statement

The authors have declared no competing interest.

### Author Declarations

United Kingdom National Research Ethics Service (11/NW/0382) gave ethical approval for this work

