## Supplementary figures and tables for "Three-dimensional motion-capture of the heart uncovers signatures of human health and disease"

### Supplementary Material

#### Supplementary Figures and Tables

##### Mass univariate analysis of signature geometry differences

Temporal signatures extracted from the latent space of the convolutional variational autoencoder were stratified into six distinct phenogroups using time series clustering. To assess localised differences in the trajectory geometry of signatures across clusters, we used a mass univariate analysis using permuted ordinary least squares (OLS) regression, as implemented in the Python module `nilearn`[1]. This method tests each feature, in this case each time point along the trajectory, independently for group-level differences, while accounting for the dependency structure through non-parametric permutation testing. This allows to avoid strong assumptions about the distribution of residuals. Formally, for each time point $t$, a linear model of the form

$$y_t = X\beta_t + \epsilon_t$$

was fitted, where $y_t$ is the temporal signature value at time $t$, $X$ is the matrix encoding phenogroup attribution, $\beta_t$ is the coefficient vector, and $\epsilon_t$ is the residual error. The significance of the coefficients was assessed through $n = 10{,}000$ permutations of the group labels, giving empirical null distributions and corrected P-values.

This analysis was performed for each pairwise combination of phenogroups and separately for the trajectories projected onto the first two principal components (PC1 and PC2), which captured the dominant axes of temporal variation. All resulting P-values were corrected for multiple comparisons using the Benjamini-Hochberg procedure, with a significance level threshold of $P < 0.05$.

As shown in Supplementary Figure 6, we observed statistically significant differences at all trajectory time points for every pair of phenogroups. However, the specific components (PC1 vs. PC2) showing differences varied across comparisons. For example, PG1 was most similar to PG3, consistent with their overlapping average trajectory shapes. Conversely, PG4 exhibited marked dissimilarity from all other phenogroups, particularly PG6, while PG2 and PG5 shared a relatively larger number of similar trajectory coordinates. Interestingly, PG6 differed substantially at the individual level from PG5, despite a similar average trajectory. This could reflect greater intra-cluster heterogeneity driven by the inclusion of extreme phenotypes: PG5 represented high-risk individuals with clinical events, while PG6 gathered an even higher-risk subgroup, potentially introducing more variation in latent temporal features.

| Number of clusters | 2 | **3** | 4 | 5 |
|---|---|---|---|---|
| Fold 1 | 0.949 | 0.971 | 0.513 | 0.656 |
| Fold 2 | 0.982 | 0.822 | 0.651 | 0.601 |
| Fold 3 | 0.875 | 0.940 | 0.733 | 0.819 |
| Fold 4 | 0.666 | 0.968 | 0.895 | 0.645 |
| Fold 5 | 0.863 | 0.869 | 0.411 | 0.861 |
| Average | 0.867 | **0.914** | 0.641 | 0.716 |

**Supplementary Table 1. Average rand index score for clustering on in-plane position.** Stability of the clustering method based on in-plane position was assessed with an average rand index score for multiple chosen values of clusters and 5 partitions of equal size of the initial dataset.

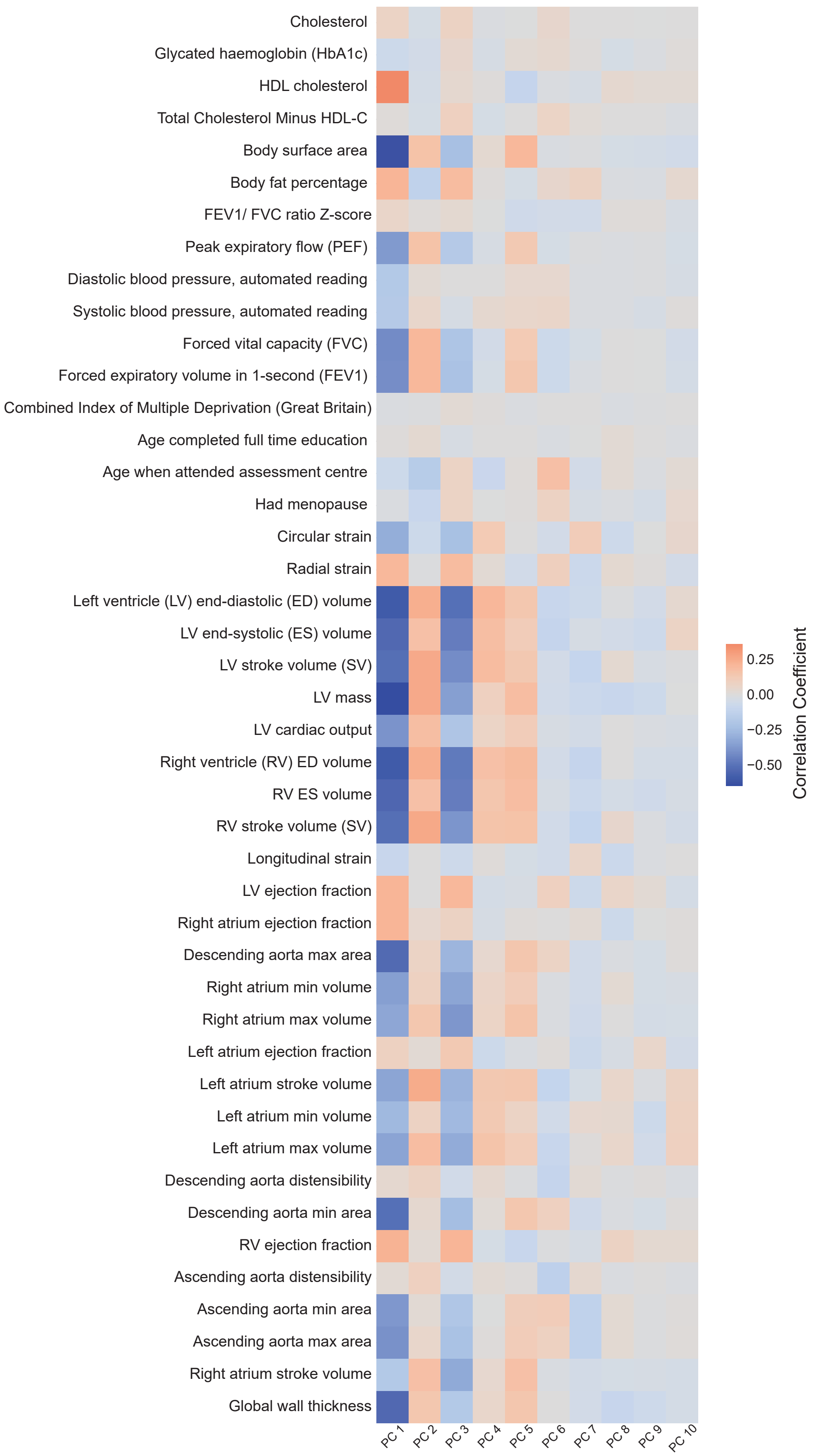


**Supplementary Figure 1. Correlation between latent principal components and clinical features.** A correlation analysis was performed between the first ten principal components of the latent space and relevant subject feature categories: demographics, body and clinical measurements, biomarkers, and cardiac MRI-derived measures.

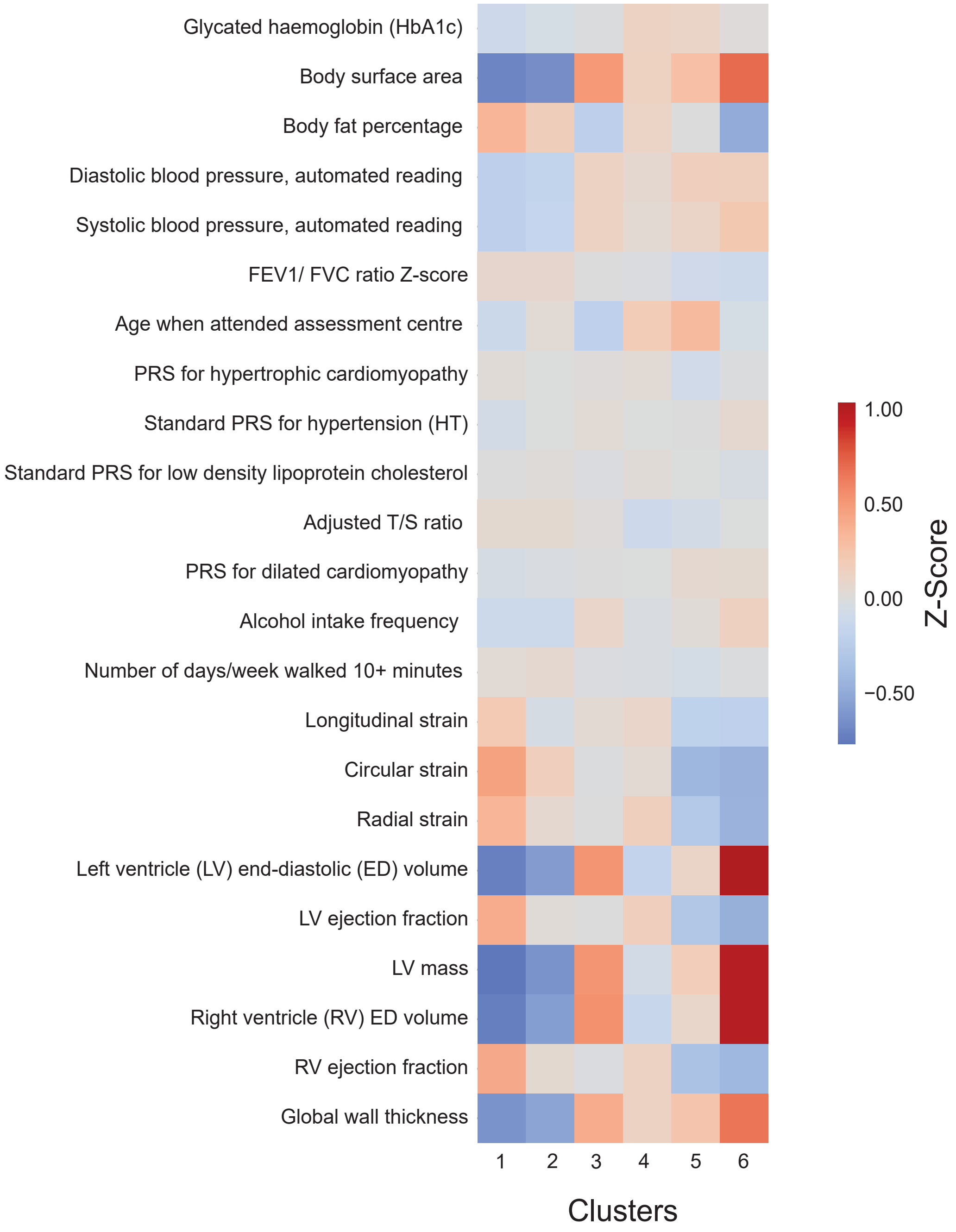


**Supplementary Figure 2. Heatmap of z-scores of important features against clusters.** Z-scores of each relevant feature are plotted for each of the six clusters. HbA1c, hemoglobin A1C; FEV1/FVC, ratio of the forced expiratory volume in 1-second to the forced vital capacity; T/S, telomere to single copy gene; PRS, polygenic risk score

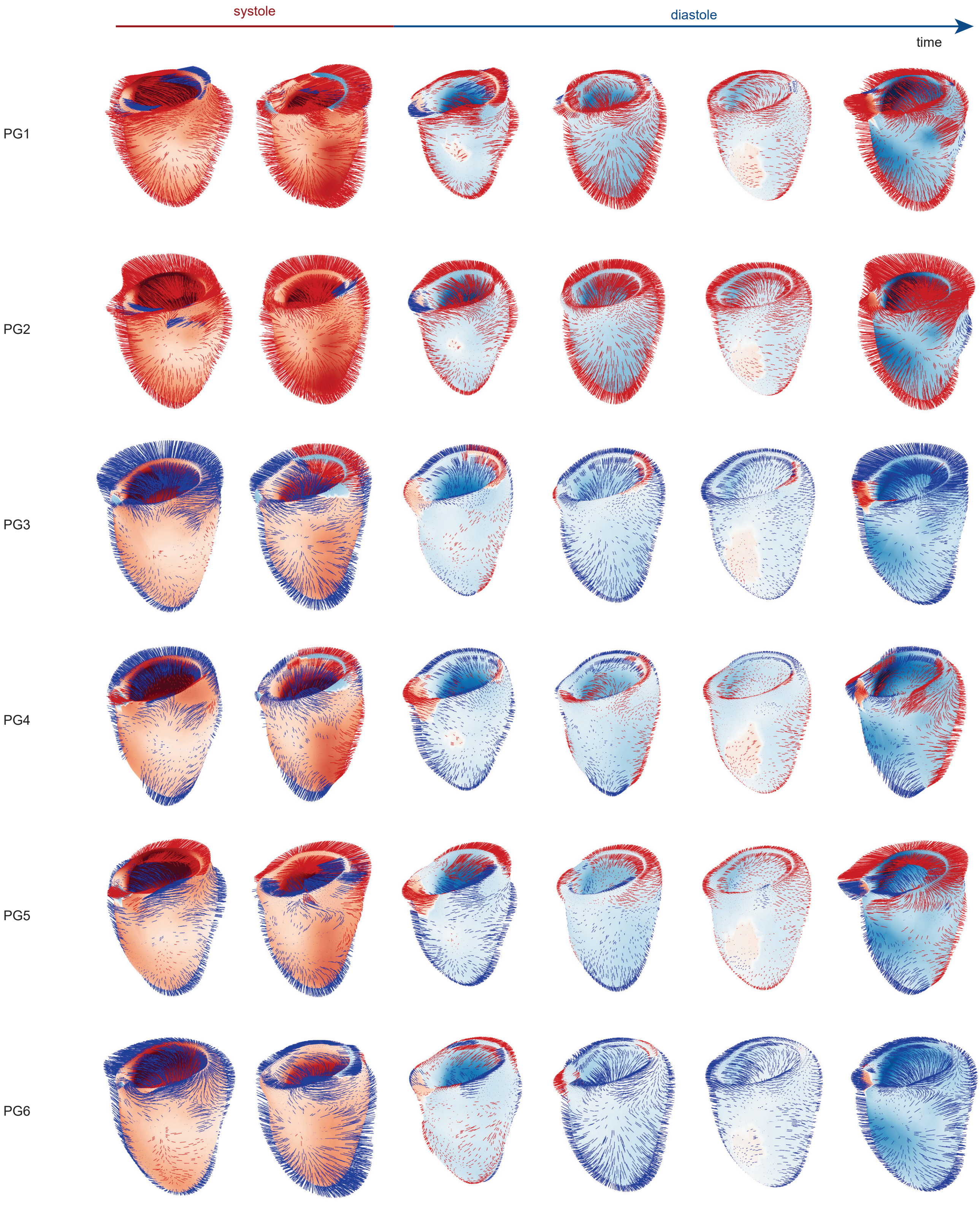


**Supplementary Figure 3. Left ventricle motion velocity by phenogroup - Anteroseptal view.** The left ventricle motion was compared to the population average motion for each phenogroup. The six frames represent one entire cardiac cycle from systole (contraction of the ventricle) to diastole (relaxation of the ventricle). The velocity of the ventricle was calculated as the local gradient of change between two consecutive frames, averaged out over multiple frames to reduce the number of displayed frames, and normalised for the size of the ventricle. Red beams indicate areas of faster movement relative to the population average motion, while blue beams show areas of relative slower movement.

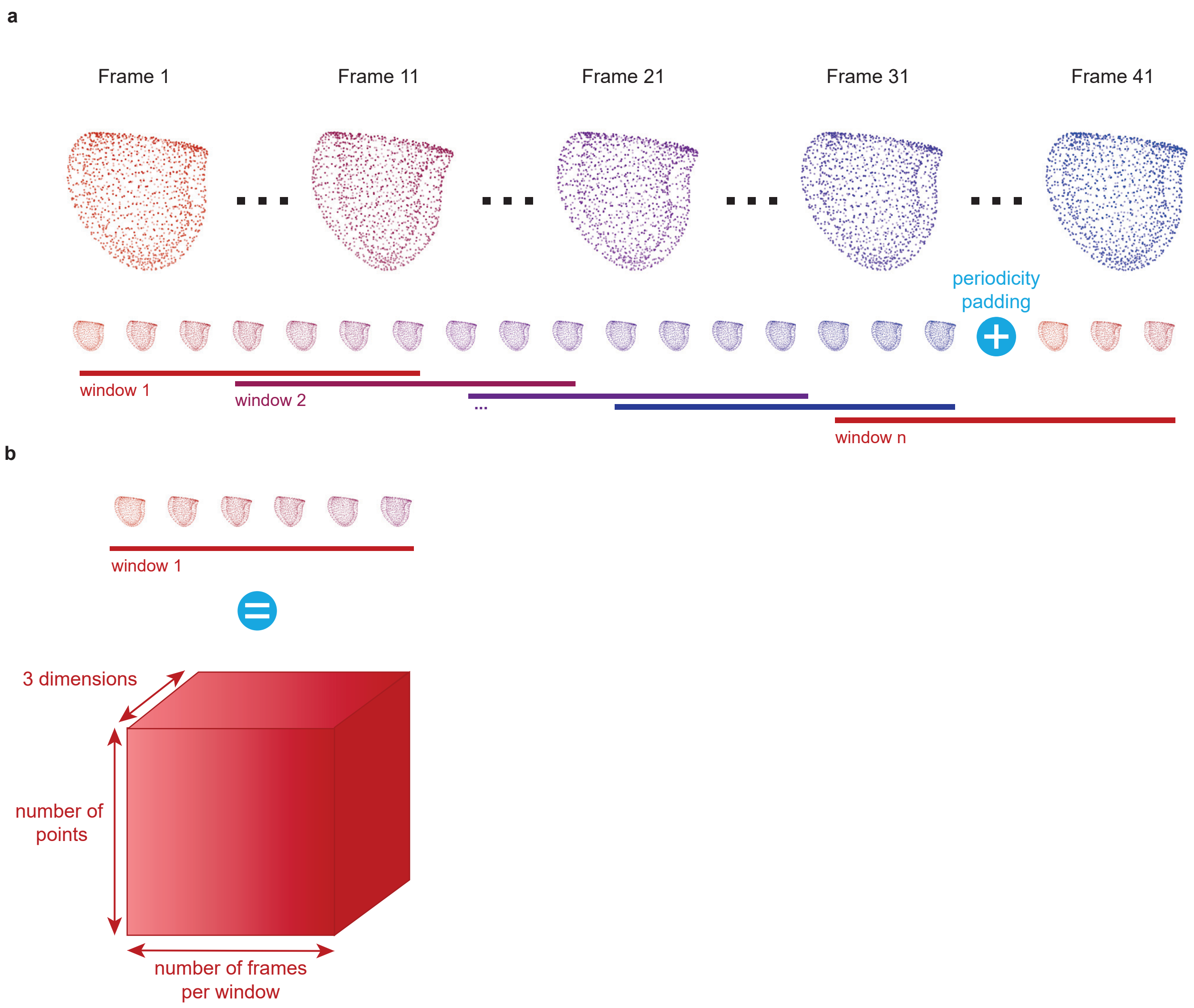


**Supplementary Figure 4. Windows of cardiac motion. a.** Windows of cardiac motion were created with a sliding window technique to embed the motion of the cardiac cycle in a single object. Periodicity padding was used to ensure that windows covered the entire cycle. **b.** Time windows were equivalent to a three-dimensional matrix in the mathematical space.

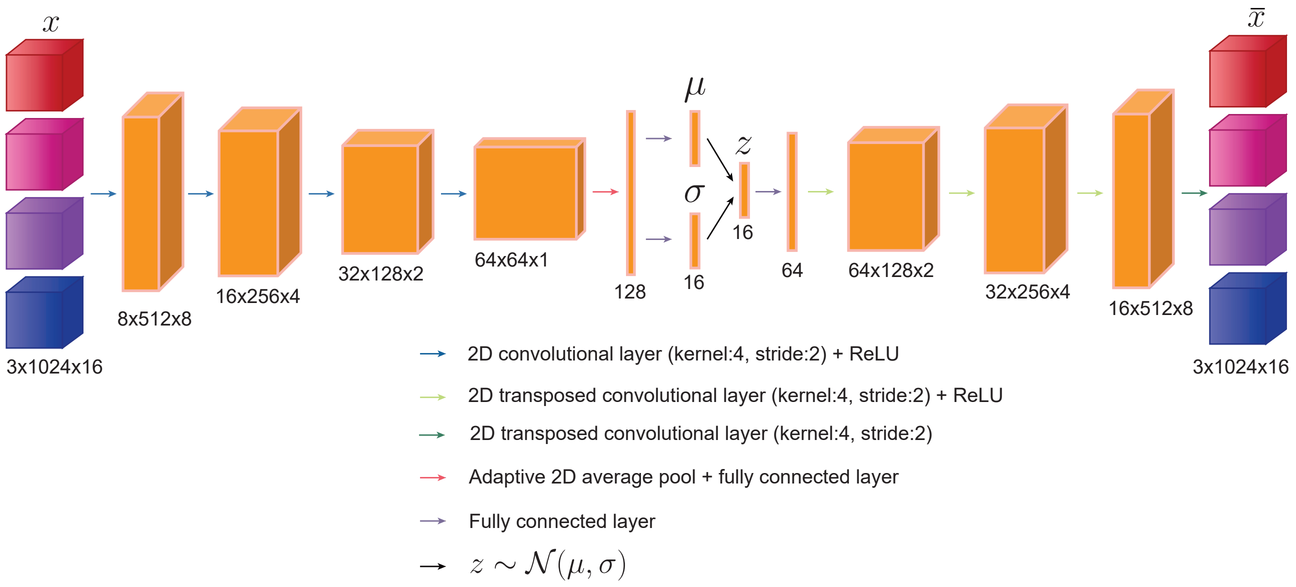


**Supplementary Figure 5. Architecture of convolutional variational autoencoder.** The convolutional autoencoder took the time windows $x$ as input. Its encoder was made of four 2D convolutional layers all followed by a reLU (rectified linear unit) activation function. Before the bottleneck, adaptive 2D average pooling was applied to the output of the convolutional layers. The latent vector $z$ was sampled from the latent distribution of mean $\mu$ and standard deviation $\sigma$. The latent vector was fed into the decoder with four 2D transposed convolutional layers, all followed by a reLU activation function except for the last layer. The output of the model was the reconstructed time windows $\bar{x}$.

| Number of clusters | **2** | 3 | 4 | 5 |
|---|---|---|---|---|
| Fold 1 | 0.937 | 0.837 | 0.748 | 0.770 |
| Fold 2 | 0.839 | 0.625 | 0.458 | 0.852 |
| Fold 3 | 0.872 | 0.400 | 0.691 | 0.683 |
| Fold 4 | 0.940 | 0.593 | 0.837 | 0.735 |
| Fold 5 | 0.932 | 0.707 | 0.296 | 0.556 |
| Average | **0.904** | 0.632 | 0.606 | 0.719 |

**Supplementary Table 2. Average rand index score for clustering on contour characteristics.** The stability of the clustering method based on contour characteristics was assessed with an average rand index score for multiple chosen values of clusters and 5 partitions of equal size of the initial dataset.

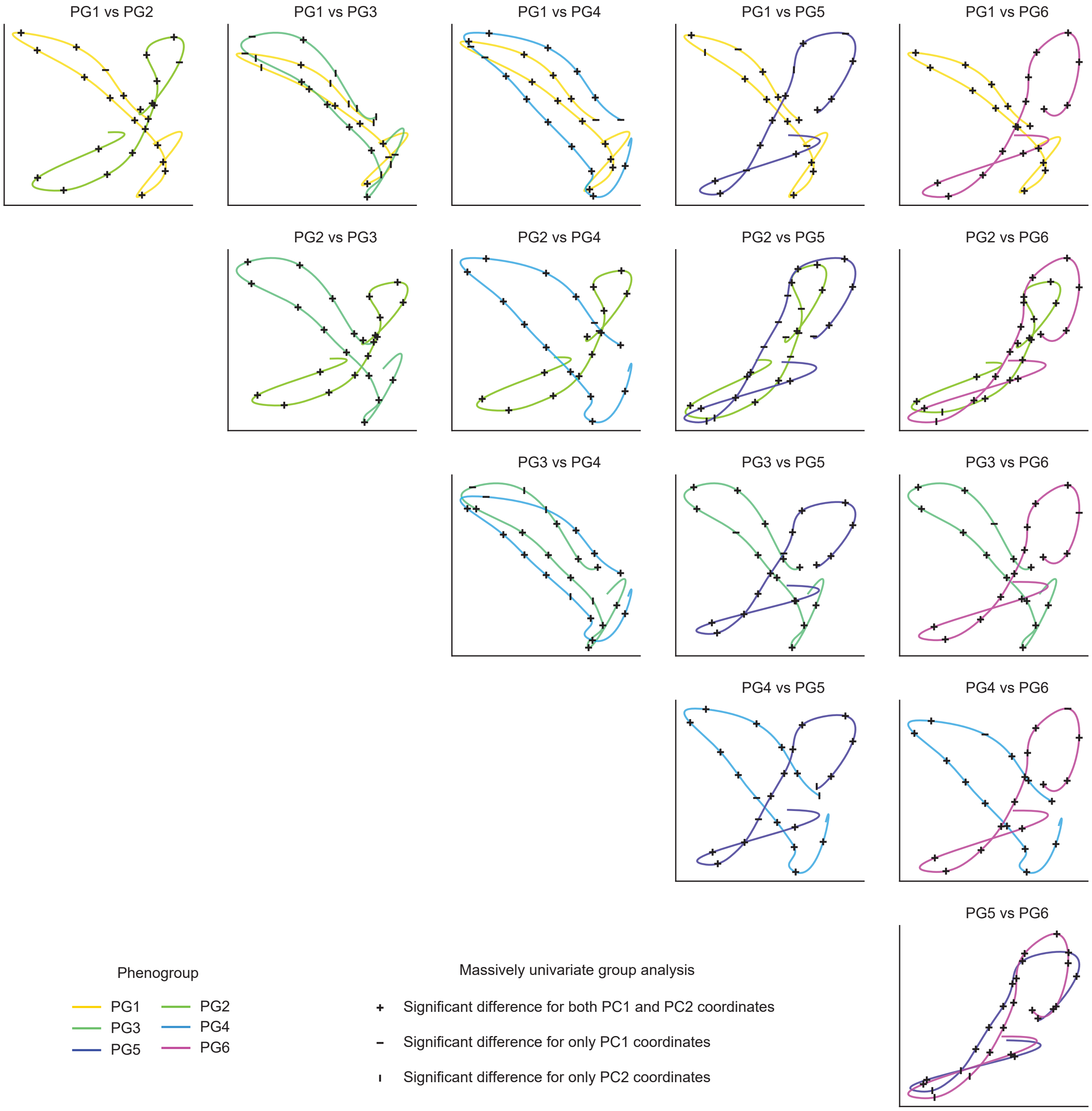


**Supplementary Figure 6. Mass univariate group analysis of geometry differences of temporal signatures** A mass univariate group analysis with permuted ordinary least squares (OLS) was performed to find geometry differences of the temporal signatures between phenogroups. The coordinates of the points making the trajectory in the space of the first and second principal components (PC1 and PC2) were compared between pairs of phenogroups. Benjamini-Hochberg correction was applied to account for multiple testing. The significance level was $P < 0.05$ and the sample size was n = 20,318. Comparisons were done for PC1 and PC2 separately. The average signatures of the phenogroups were plotted, and each trajectory point was annotated by a symbol indicating a significant difference between PC1, PC2, or both for the pair of phenogroups.

| Image-derived measure | Unit |
|---|---|
| Longitudinal strain | % |
| Circumferential strain | % |
| Radial strain | % |
| LV end-diastolic volume | mL |
| LV end-systolic volume | mL |
| LV stroke volume | mL per beat |
| LV cardiac output | mL per minute |
| LV ejection fraction | % |
| LV mass | mg |
| Global wall thickness | mm |
| RV end-diastolic volume | mL |
| LV end-systolic volume | mL |
| RV stroke volume | mL per beat |
| RV ejection fraction | % |
| Ascending aorta max area | mm2 |
| Ascending aorta min area | mm2 |
| Ascending aorta distensibility | $10^3$ $mmHg^{-1}$ |
| Descending aorta max area | mm2 |
| Descending aorta min area | mm2 |
| Descending aorta distensibility | $10^3$ $mmHg^{-1}$ |
| Left atrium max volume | mL |
| Left atrium min volume | mL |
| Left atrium stroke volume | mL per beat |
| Left atrium ejection fraction | % |
| Right atrium max volume | mL |
| Right atrium min volume | mL |
| Right atrium stroke volume | mL per beat |
| Right atrium ejection fraction | % |

**Supplementary Table 3. Image-derived measures for benchmark data.** We used the following MRI-derived clinical measurements in place of the spatiotemporal cardiac motion data to assess its discriminative power.

| | Data | Model | Clustering | Stability (ARI) |
|---|---|---|---|---|
| Our model | High-dimensional meshes | CVAE+PCA Non-linear | Time series K-means | In-plane position: 0.91<br>Contour: 0.90 |
| Benchmark data | Tabular MRI-derived measures | UMAP+PCA Non-linear | K-means | 0.82 |
| Benchmark model | High-dimensional meshes | PCA Linear | Time series K-means | In-plane position: 0.50<br>Contour: 0.85 |

**Supplementary Table 4. Comparison of our model to two benchmarks.** The benchmark against our data substituted cardiac point clouds with tabular MRI-derived data. The benchmark against our model used a simple principal component analysis (PCA) applied to the windows of cardiac motion. Our model achieved better cluster stability and robustness. ARI, average Rand index score; CVAE, convolutional variational autoencoder; UMAP, uniform manifold approximation and projection.

| Phenotype | ICD 10 | ICD 10 String | Phenocode |
|---|---|---|---|
| Atherosclerosis | I25.0 | Atherosclerotic cardiovascular disease, so described | 440 |
| Atherosclerosis | I70 | Atherosclerosis | 440 |
| Atherosclerosis | I70.8 | Atherosclerosis of other arteries | 440 |
| Atherosclerosis | I70.9 | Generalized and unspecified atherosclerosis | 440 |
| Cardiac arrest | I46 | Cardiac arrest | 427.42 |
| Cardiac arrest | I46.0 | Cardiac arrest with successful resuscitation | 427.42 |
| Chronic pulmonary heart disease | I27.1 | Kyphoscoliotic heart disease | 415.2 |
| Chronic pulmonary heart disease | I27.8 | Other specified pulmonary heart diseases | 415.2 |
| Chronic pulmonary heart disease | I27.9 | Pulmonary heart disease, unspecified | 415.2 |
| Coronary atherosclerosis | I24.0 | Coronary thrombosis not resulting in myocardial infarction | 411.4 |
| Coronary atherosclerosis | I25.1 | Atherosclerotic heart disease | 411.4 |
| Dilated cardiomyopathy | I42.0 | Dilated cardiomyopathy | 425.1 |
| Essential hypertension | I10 | Essential (primary) hypertension | 401.1 |
| Heart failure NOS | I50 | Heart failure | 428.2 |
| Heart failure NOS | I50.1 | Left ventricular failure | 428.2 |
| Heart failure NOS | I50.9 | Heart failure, unspecified | 428.2 |
| Hypertrophic cardiomyopathy | I42.1 | Obstructive hypertrophic cardiomyopathy | 425.11 |
| Hypertrophic cardiomyopathy | I42.2 | Other hypertrophic cardiomyopathy | 425.12 |
| Mitral valve disease | I05 | Rheumatic mitral valve diseases | 394.2 |
| Mitral valve disease | I05.0 | Mitral stenosis | 394.2 |
| Mitral valve disease | I05.1 | Rheumatic mitral insufficiency | 394.2 |
| Mitral valve disease | I05.2 | Mitral stenosis with insufficiency | 394.2 |
| Mitral valve disease | I05.8 | Other mitral valve diseases | 394.2 |
| Mitral valve disease | I05.9 | Mitral valve disease, unspecified | 394.2 |
| Mitral valve disease | I08.1 | Disorders of both mitral and tricuspid valves | 394.2 |
| Mitral valve disease | I34.0 | Mitral (valve) insufficiency | 394.2 |
| Mitral valve disease | I34.1 | Mitral (valve) prolapse | 394.2 |
| Myocardial infarction | I21 | Acute myocardial infarction | 411.2 |
| Myocardial infarction | I21.0 | Acute transmural myocardial infarction of anterior wall | 411.2 |
| Myocardial infarction | I21.1 | Acute transmural myocardial infarction of inferior wall | 411.2 |
| Myocardial infarction | I21.2 | Acute transmural myocardial infarction of other sites | 411.2 |
| Myocardial infarction | I21.3 | Acute transmural myocardial infarction of unspecified site | 411.2 |
| Myocardial infarction | I21.4 | Acute subendocardial myocardial infarction | 411.2 |
| Myocardial infarction | I21.9 | Acute myocardial infarction, unspecified | 411.2 |
| Myocardial infarction | I22 | Subsequent myocardial infarction | 411.2 |
| Myocardial infarction | I22.0 | Subsequent myocardial infarction of anterior wall | 411.2 |
| Myocardial infarction | I22.1 | Subsequent myocardial infarction of inferior wall | 411.2 |
| Myocardial infarction | I22.8 | Subsequent myocardial infarction of other sites | 411.2 |
| Myocardial infarction | I22.9 | Subsequent myocardial infarction of unspecified site | 411.2 |
| Myocardial infarction | I23 | Certain current complications following acute myocardial infarction | 411.2 |
| Myocardial infarction | I23.0 | Haemopericardium as current complication following acute myocardial infarction | 411.2 |
| Myocardial infarction | I23.1 | Atrial septal defect as current complication following acute myocardial infarction | 411.2 |
| Myocardial infarction | I23.2 | Ventricular septal defect as current complication following acute myocardial infarction | 411.2 |
| Myocardial infarction | I23.3 | Rupture of cardiac wall without haemopericardium as current complication following acute myocardial infarction | 411.2 |
| Myocardial infarction | I23.6 | Thrombosis of atrium, auricular appendage, and ventricle as current complications following acute myocardial infarction | 411.2 |
| Myocardial infarction | I23.8 | Other current complications following acute myocardial infarction | 411.2 |
| Myocardial infarction | I24.1 | Dressler syndrome | 411.2 |
| Myocardial infarction | I25.2 | Old myocardial infarction | 411.2 |
| Myocardial infarction | I51.0 | Cardiac septal defect, acquired | 411.2 |
| Myocardial infarction | I51.3 | Intracardiac thrombosis, not elsewhere classified | 411.2 |
| Other cardiomyopathy | I42 | Cardiomyopathy | 425.1 |
| Other cardiomyopathy | I42.3 | Endomyocardial (eosinophilic) disease | 425.1 |
| Other cardiomyopathy | I42.4 | Endocardial fibroelastosis | 425.1 |
| Other cardiomyopathy | I42.5 | Other restrictive cardiomyopathy | 425.1 |
| Other cardiomyopathy | I42.6 | Alcoholic cardiomyopathy | 425.8 |
| Other cardiomyopathy | I42.7 | Cardiomyopathy due to drugs and other external agents | 425 |
| Other forms of chronic heart disease | I51.1 | Rupture of chordae tendineae, not elsewhere classified | 414 |
| Other forms of chronic heart disease | I51.2 | Rupture of papillary muscle, not elsewhere classified | 414 |
| Other forms of chronic heart disease | I51.5 | Myocardial degeneration | 414 |
| Other forms of chronic heart disease | I51.8 | Other ill-defined heart diseases | 414 |
| Other forms of chronic heart disease | I51.9 | Heart disease, unspecified | 414 |
| Other forms of chronic heart disease | I52 | Other heart disorders in diseases classified elsewhere | 414 |

**Supplementary Table 5. Phenotypes and associated ICD-10 codes.** Cardiovascular diseases and outcomes were grouped under related phenotypes for subsequent analysis.
